# Measuring the Quality of AI-Generated Clinical Notes: A Systematic Review and Experimental Benchmark of Evaluation Methods

**DOI:** 10.1101/2025.11.18.25340507

**Authors:** Alexandra Dahlberg, Tiila Käenniemi, Tiia Winther-Jensen, Olli Tapiola, Rami Luisto, Tuukka Puranen, Max Gordon, Enni Sanmark, Ville Vartiainen

**Affiliations:** Clinicum, Faculty of Medicine, University of Helsinki, Helsinki, Finland; Mehiläinen, Helsinki, Finland; Faculty of Arts, University of Helsinki, Finland; Gofore Lead Oy, Helsinki, Finland; Faculty of Information Technology, University of Jyväskylä, Finland; Digital Workforce Services Plc, Helsinki, Finland; Department of Clinical Sciences, Danderyd Hospital, Karolinska Institutet, Stockholm, Sweden; Heart and Lung Center, Helsinki University Hospital, Helsinki, Finland

**Author notes:** Contributed equally (Tiila Käenniemi, Tiia Winther-Jensen).

## Abstract

**Background:** High-quality clinical documentation is essential for safe, effective care, yet producing it is time consuming and error prone. Large language models (LLMs) can assist with note generation, but clinical adoption is determined by the resulting note quality. However current evaluation practices vary, and their clinical relevance is unclear. Drawing on a multidisciplinary perspective, we examined how quality is assessed and how those assessments align with clinical demands.

**Methods:** We systematically searched Ovid Medline and Scopus on 10 April 2025 for peer-reviewed studies that used LLMs in generating clinical notes and included an evaluation of the quality of the resulting text. The screening followed PRISMA and the protocol was preregistered in PROSPERO. Data on metrics, and outcomes were synthesised narratively. Based on these findings, we designed an experimental setup to test the most common evaluation metrics and an LLM-as-evaluator, included for its scalability across large test sets. The experiment used synthetic cases with targeted perturbations.

**Findings:** Thirty-seven studies were included. The reporting was dominated by lexical overlap metrics, chiefly ROUGE and BLEU. Semantic similarity metrics, such as BERTScore and BLEURT, were less common. A human evaluation was frequent but heterogeneous, with criteria and methods defined using varying degrees of detail; the most common foci were correctness, fluency, and aspects of clinical acceptability. In our experimental setup, lexical overlap metrics detected deletions and modifications but penalised meaning-preserving paraphrases. Semantic metrics and LLM-as-evaluators were more tolerant of paraphrased perturbations, yet remained sensitive to relevant changes, with performance varying by model and language.

**Interpretation:** Current practice relies on lexical overlap metrics that are useful for cursory checks but insufficient as proxies for quality. We recommend a layered strategy that pairs semantic metrics with LLM-as-evaluator for scalability and includes targeted human adjudication. Broader, safety-focused validation across institutions and languages is needed before routine deployment.

**Funding:** Business Finland through the GenAID research project. Personal grants listed under Acknowledgments.

## Introduction

High-quality clinical documentation is integral to safe and effective care. Clear, complete notes support decision making, reduce treatment errors, and maintain continuity, whereas incomplete, disorganised, or inaccurate records result in miscommunication, delay diagnosis, and worsen outcomes^1,2^. Interventions including dictation, structured templates, and medical scribes provide partial mitigation, but evidence for sustained improvement is mixed and quality remains variable^1,2^. Recent advances in artificial intelligence (AI) have renewed the interest in assistance for clinical documentation. Large language models (LLMs) are AI systems capable of large variety of natural language processing tasks, enabling human-like text generation and cross-task generalisation^3^. LLMs can draft histories, discharge summaries and other clinical notes from transcripts or structured data. However, during the process LLMs transform content and may omit critical details or introduce misinformation underscoring the need for robust evaluation^2^. Increasing the degree of automation and scaling LLM use requires automated and scalable evaluation methods that account for these effects of the generation process.

To assess the current evaluation practices, we first conducted a systematic review of peer-reviewed studies evaluating AI-generated clinical notes, cataloguing the metrics, and criteria employed. We then implemented a complementary experimental framework that tested the widely used evaluation methods on synthetic clinical cases with controlled perturbations: deletions, modifications, and paraphrases. In contrast to earlier reviews that emphasise checklist-style content audits, our focus is the quality of AI-generated text and its evaluation, with the aim of identifying which assessment approaches best reflect clinical utility and safety and can be automated at scale.

## Methods

This study has two components. First, we conducted a systematic review of peer-reviewed studies that reported an evaluation of AI-generated clinical notes in accordance with PRISMA guidelines. Second, an experimental framework that operationalises several evaluation criteria identified in the review. Using synthetic clinical cases with targeted perturbations, we evaluate standard automated metrics alongside several LLM-based evaluators, comparing performance across languages and perturbation types. This design lets the experiment probe, in a controlled setting, the strengths and limitations highlighted by the review.

### Search strategy and selection criteria

This systematic review was conducted in accordance with the PRISMA guidelines^4^ (Supplementary Table S1). The review protocol was preregistered at PROSPERO (CRD420251044770). Ovid MEDLINE and Scopus were searched on 10 April 2025. Search terms captured concepts of artificial intelligence and clinical documentation quality. Strings combined keywords and MeSH terms including, but not limited to, clinical note, AI, LLM, RAG, and evaluation. Full syntaxes for each database are provided in Supplementary Table S2. No date limits were applied. Criteria for inclusion and exclusion are presented in Supplementary Table S3.

After removal of duplicates, the titles and abstracts were screened in parallel by two reviewers working as a pair, with a third reviewer independently assessing all records. Discrepancies were resolved by discussion. Full texts were then assessed and data extracted using the same process. The PRISMA flow diagram summarises the process (Figure 1). In total, 37 studies met the inclusion criteria and were included in the review.

**Figure 1.**
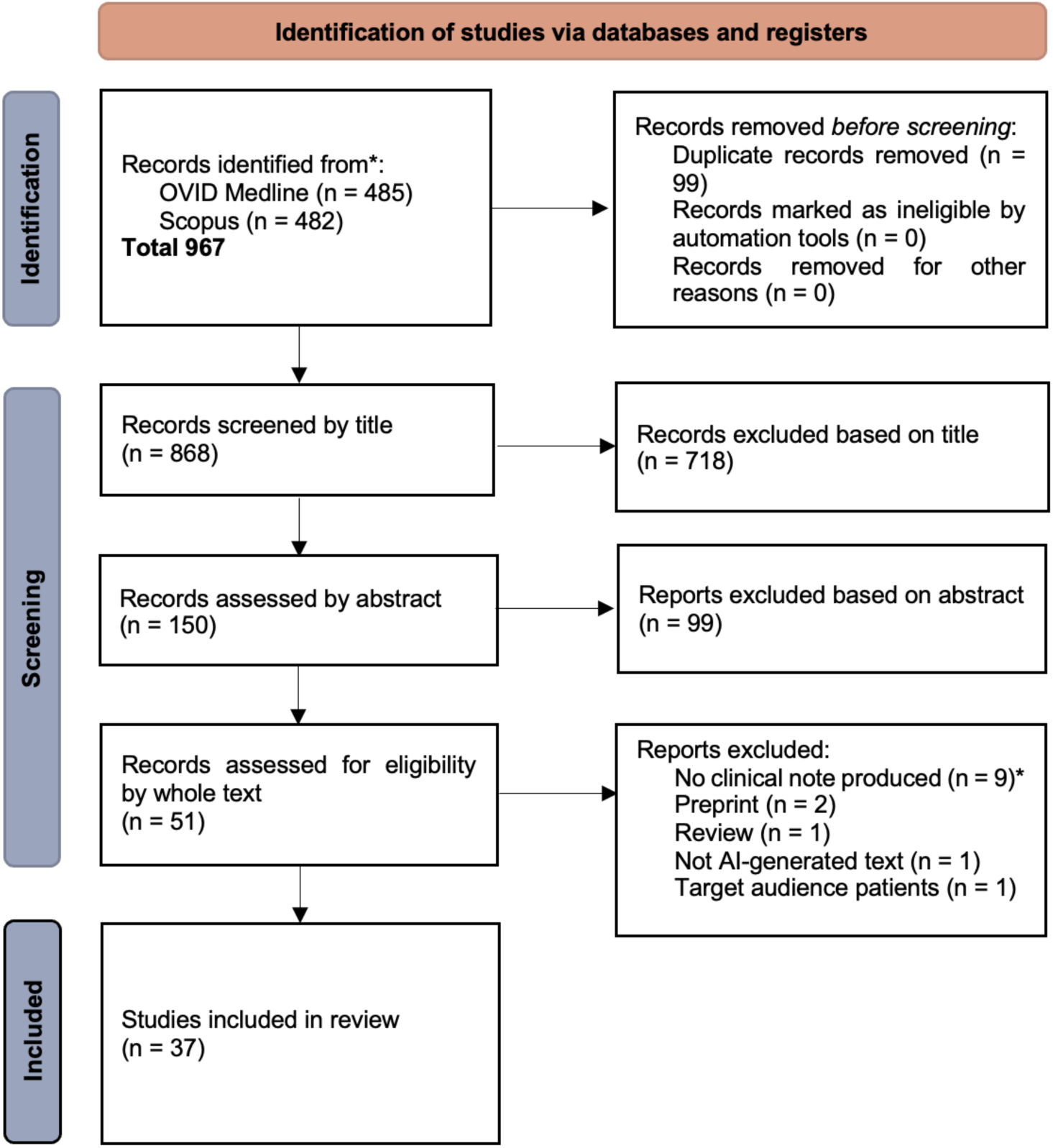
PRISMA flow diagram^4.^ *Records excluded as “no clinical note produced” generated Q&A content, diagnosis lists, or image categorization instead of documentation aimed at healthcare personnel.

#### Data Extraction and Quality Assessment

Data were extracted using a standardised form capturing publication details (authors, journal, year), clinical setting or context (specialty, use case for AI text generation), AI system or model type (for example, GPT-3 (proprietary)), and the methods used to evaluate the generated text quality. For evaluation methods, we recorded automated metrics, the extent and means of human evaluation, the criteria, and clinical standard checklists used, and any use of large language models as evaluators. Where available, we captured quantitative results, for example means and statistical significance, and qualitative findings, for example narrative comments on note quality.

Because study designs varied, from randomised controlled trials to observational usability studies, we qualitatively considered methodological rigour, for example randomisation, blinding of evaluators, and sample size, but did not exclude studies based on quality and did not perform a formal risk-of-bias scoring.

#### Synthesis of Results

We synthesised findings narratively, grouped by themes in evaluation approach and outcome. Interventions, comparators, and outcome measures were too heterogeneous for meta-analysis, and no common effect measures, such as risk ratios or mean differences, were applicable. For each thematic synthesis we first tabulated study characteristics against our planned categories, text-to-text, image-to-text, and evaluation methodology. Text-to-text denotes systems that generate text from textual inputs; image-to-text denotes multimodal systems that produce text from visual inputs. The evaluation methodology category comprises studies whose primary contribution is the design, comparison, or validation of evaluation methods rather than the assessment of a specific AI-generated clinical note, typically to benchmark a newly proposed metric. Using this scheme, 29 studies addressed text-to-text generation, 5 addressed image-to-text generation, and 3 focused on evaluation methodology.

Evaluation approaches were divided into two broad categories: automated and human evaluation. The synthesis of automated approaches focused on the reported metrics, and the synthesis of human evaluation on harmonization of criteria. Reporting of human evaluation approaches, terminology and definitions for quality criteria varied across papers and we harmonised terminology into correctness, completeness, conciseness, fluency, clinical acceptability, and other criteria using explicit mapping rules. The preferred criteria were selected for their recurrence and conceptual coherence across the reviewed literature. These criteria represent frequently used yet inconsistently labelled or defined constructs across studies; grouping them under broader, conceptually coherent terms enables synthesis of results, highlights the lack of standardization in evaluation terminology, and allows for future refinement and subcategorization. The terms correctness, completeness, conciseness, and fluency were chosen for their high frequency of occurrence and conceptual consistency across studies. The term clinical acceptability was selected to encompass the multidimensional nature of clinical suitability, including safety, usefulness, professional trust, and practical applicability. It was preferred over narrower alternatives such as clinical validity, utility, or appropriateness because it integrates these aspects, enabling both consistent synthesis and future analysis of the dimensions and constituents of clinical acceptability. Other criteria were included to capture evaluation aspects that could not be mapped elsewhere due to insufficient specificity of the reported information.

Reported percentages were converted to n/N_x_ where denominators were available. If a metric variant was unspecified, for example “ROUGE” with no variant, we recorded it as unspecified and flagged it in the tables. When required information was missing or unclear, we coded it as ‘not reported’.

### Design of the Experimental Setup

To assess the selected automated evaluation tools, we used synthetic cases from MedBench with permission. MedBench is a multilingual, multi-specialty benchmark in development, comprising fictional clinical notes with an associated discharge summary for each patient; available to registered users online^5^. From this resource, we selected all five internal medicine discharge summaries available in English, Swedish and Finnish. We then constructed perturbations ourselves, covering three edit types: (1) deletion of clinically relevant information, (2) modification of key details, and (3) paraphrase, for example via synonym substitution. Each type was applied at three intensities, approximately five, ten and fifteen edits, yielding nine perturbated variants per case. Edit schemas were held consistent across languages. All clinical notes and perturbations in all three languages are available in the repository cited in the Data Sharing statement.

#### Metrics

Guided by the systematic review, we evaluated four families of automated metrics. Foremost, we considered lexical overlap metrics, namely BLEU-1 to BLEU-4, and ROUGE-1, ROUGE-2 and ROUGE-L, which compare exact tokens or n-grams between a system output and its reference, and therefore capture surface similarity rather than meaning. Subsequently, we considered semantic similarity metrics, which map texts to vector representations and score their proximity in order to capture meaning beyond wording. This family includes embedding-based measures (for example BERTScore), learned reference-based metrics (BLEURT), and model-based likelihood metrics (BARTScore). In our experiments we used BLEURT and BERTScore with RoBERTa-large and XLM-R-base backbones. We also included a text-embedding cosine similarity on theoretical grounds as a direct estimate of semantic proximity. Although not reported in the review, many semantic metrics rely on cosine similarity in the scoring step. Embeddings were computed using text-embedding-3-large (OpenAI, San Francisco, USA) embedding model. Full implementation details, including prompts, checkpoints and the repository, are provided in the Data Availability statement to ensure variant consistency across languages. Finally, to examine the LLM-as-evaluator paradigm, we prompted a set of LLMs to rate the divergence between each system output and its reference. The model set comprised Gemma 3n E2B, Gemma 3n E4B, DeepSeek R1, gpt-oss-20B, gpt-oss-120B, GPT-3.5, GPT-4, GPT-4o, GPT-4.1, GPT-5 mini, GPT-5 nano and GPT-5. All model runs used a fixed decoding temperature and maximum output tokens as specified in the repository. For the open-source 20B model, a 4000-token limit yielded scores for 78 of 135 edited inputs on the first pass, with reruns increasing coverage to 89 of 135; increasing the limit to 5000 tokens produced 89 of 135 on the first pass. For the 120B model, a 4000-token limit produced scores for all but two inputs on the first pass, and reruns achieved complete coverage. All configurations, prompts, and model hashes are versioned for reproducibility. Python version (Python Software Foundation, Wilmington, USA)^6–15^.

#### Statistics

For each metric–model pair, we computed summary scores within each language (English, Swedish, Finnish) by perturbation type (delete, modify, paraphrase) and perturbation level (1-3) by comparison to the original summary of the medical case.

To assess the association between perturbation level and evaluation metric scores we used linear mixed-effects model with perturbation level implemented as continuous variable and case as random effect. Graphical inspection indicated that the categories behaved approximately linearly with level, which justifies the chosen approach.

No ethical approval was required as the experiment used synthetic clinical notes and involved neither human participants nor protected health information.

## Results

### Systematic Review

We synthesised the methods used to assess the quality of AI-generated clinical text in the literature. All included studies are listed in Supplementary Table S4.

#### Lexical Overlap Metrics Dominate Current Practice

In current research addressing clinical note generation, lexical overlap metrics regarding lexical and syntactic characteristics, chiefly ROUGE^16^ and BLEU)^17^, remain dominant (Table 1).

**Table 1.** Automated metrics used in included studies. Values are n/N (%). Denominators for percentage counts: text-to-text n_t_=29, image-to-text n_i_=5, evaluation methodology n_e_=3, total N=37. A study is counted if it reported any score for the metric, regardless of variant. Full study-to-metric mappings are provided in Supplementary Table S5.

| **Automated Metric** | **Text-to-text (n)** | **Image-to-text (n)** | **Evaluation methodology (n)** |
| --- | --- | --- | --- |
| BLEU-1 | 0·0% (0) | 40·0% (2) | 0·0% (0) |
| BLEU-2 | 0·0% (0) | 40·0% (2) | 0·0% (0) |
| BLEU-3 | 0·0% (0) | 40·0% (2) | 0·0% (0) |
| BLEU-4 | 3·4% (1) | 60·0% (3) | 0·0% (0) |
| BLEU | 6·9% (2) | 20·0% (1) | 0·0% (0) |
| ROUGE-1 | 51·7% (15) | 20·0% (1) | 33·3% (1) |
| ROUGE-2 | 48·3% (14) | 20·0% (1) | 33·3% (1) |
| ROUGE-3 | 6·9% (2) | 0·0% (0) | 0·0% (0) |
| ROUGE-4 | 3·4% (1) | 0·0% (0) | 0·0% (0) |
| ROUGE-L | 62·1% (18) | 100·0% (5) | 33·3% (1) |
| ROUGE-Lsum | 10·3% (3) | 0·0% (0) | 33·3% (1) |
| BERTScore<sup>13</sup> | 41·4% (12) | 40·0% (2) | 100·0% (3) |
| BLEURT<sup>18</sup> | 13·8% (4) | 0·0% (0) | 33·3% (1) |
| BARTScore<sup>19</sup> | 0·0% (0) | 0·0% (0) | 66·7% (2) |

In addition to more widely used BLEU and ROUGE families, two text-to-text and one image-to-text studies used METEOR<sup>20</sup> and one image-to-text study used CIDEr<sup>21</sup>. Both measures incorporate semantic approaches offering modest improvements to BLEU and ROUGE families.<sup>21</sup>

#### Growing Interest in Semantic Metrics

As shown in Table 1, BERTScore (backbone BERT^22^) was the most commonly applied semantic metric in our review, followed by BLEURT^18^ (BERT^22^ and RemBERT) and BARTScore^19^ (backbone BART).

In parallel, more domain-adapted semantic evaluation approaches have been proposed to better reflect clinical accuracy and relevance. By domain-adapted, we mean metrics specifically designed for e.g. medical use. In our review these evaluations were not uncommon, but the use of individual metrics was highly fragmented and often limited to single studies. Abacha et al. (2023)^23^ used medically weighted models (e.g., MedBERTScore^23^, MedBARTScore^23^, ClinicalBLEURT^23^) which consistently outperformed their non-domain-adapted counterparts. Other domain-adapted semantic metrics mentioned only once were SummaC^24^, entailment-based CTC (Compression, Transduction, and Creation)^25^, and Dependency Arc Entailment (DAE)^26^.

#### Medical Concept and Factuality Metrics

In addition to domain-adapted varieties of lexical overlap and semantic metrics, specialized metrics have also been proposed to account for highly specialized medical vocabulary and factual correctness.

The MIST (Medical Information Semantic Textual similarity)^27^ metric was reported in one study. It uses knowledge-graph embeddings derived from the Unified Medical Language System (UMLS)^28^ to improve concept-level similarity assessment, particularly for rare terms and factual statements in generated versus reference notes.

Other domain-adapted metrics, each reported once, included AlignScore^29^, FactScore^30^, and F1-score based on RadGraph^31^, and UMLS^28^ databases, each targeting aspects of factuality, structured information extraction, or concept coverage. For instance, F1-RadGraph quantifies the overlap of radiology-specific entities and relations, while FactScore measures factual consistency against verified medical statements.

#### LLM-Based Evaluation Trends

LLM-as-evaluator was a rare approach in the included literature. Three studies used LLMs directly to appraise AI-generated notes. Brake and Schaaf (2024)^32^ tested LLaMA-2 (7B, 13B, 70B) for factual consistency in PEGASUS-generated SOAP (Subjective, Objective, Assessment and Plan) notes, comparing ROUGE and factuality scores with human review of 40 of 543 vignettes and LLM review under zero-to three-shot prompting. Agreement between humans and LLaMA-2-70B (two-shot) was substantial for age (κ = 0.79), perfect for gender (κ = 1.00), and fair for body part (κ = 0.32), indicating that LLaMA-2 may reliably reproduce human annotations for some narrowly defined attributes, while its performance and therefore, potential generalisability, remains uncertain for more complex or less consistently annotated categories. Schumacher et al. (2024)^33^ introduced MED-OMIT (GPT-4), an omission-oriented metric that counts reference facts excluded from summaries and weights them by clinical relevance, benchmarking it against established metrics. It showed broad agreement with clinical experts on both omission detection and importance, with weak or nonsignificant correlations with ROUGE and BERTScore for stronger models. Mishra et al. (2024)^34^ proposed G-Eval (GPT-4) to generate synthetic edit feedback, providing targeted revision instructions to improve outputs from smaller models, fewer than 10B parameters. Clough et al. (2024)^35^ applied an OpenAI text classifier to AI-generated discharge summaries and found all labelled “very unlikely” to be AI-generated, highlighting current limits of detection tools.

#### Human Evaluation and Evaluation Criteria

Human evaluation featured in nearly all reviewed articles, although implementation varied (Supplementary Table S6). It was applied systematically in 27 of the 37 studies. Sampling strategies varied: 12 studies evaluated fewer than half of all vignettes, whereas 16 assessed more than 50 per cent. A few studies used a single evaluator per note, most used at least three, and the maximum observed was five. Evaluators were typically medical professionals or students with heterogeneous levels of experience. Few studies reported inter-rater reliability, and reporting of other statistic was uncommon.

Due to the heterogeneous nature of human evaluation approaches in the reviewed papers, human evaluation findings are reported according to the assessed quality criteria. Our harmonized categories for criteria along with concise definitions and examples are given in Supplementary Table S7. Using the harmonized categories, we quantified how often each criterion appeared in the included studies, by study category.

The most commonly used criterion was correctness (text-to-text 62% [18], image-to-text 60% [3], evaluation methodology 100% [3]), typically defined as the inclusion of accurate information and the absence of fabricated or incorrect statements. Completeness (52% [15], 40% [2], 100% [3]) was the next most frequent, usually operationalised as coverage of, or omission of, required data elements against a predefined checklist. Fluency (45% [13], 20% [1], 0% [0]) was commonly rated on Likert scales, although rubrics were rarely standardised.

Clinical acceptability (55% [16], 60% [3], 0% [0]) spanned PDQI-9^36^ dimensions such as organisation, usefulness, and timeliness, as well as constructs including clinical validity, perceived harm or likelihood of harm, appropriateness to context, willingness to use the note in practice, time efficiency, consultation quality, and practical validation in a hospital setting. Conciseness (21% [6], 0% [0], 0% [0]) was assessed less often, typically in terms of output length or redundancy. Less common criteria were grouped under other criteria (31% [9], 0% [0], 33% [1]); these included, for example, distinguishability of AI-generated from human-written text and aspects of structure and formatting. Criteria that lacked sufficient specificity to be mapped to a single category, such as intuitive global judgements of quality, were also assigned to this group. Where appropriate, a single assessment was recorded under multiple categories; for instance, “inclusion of key clinical parameters” was coded as both completeness and clinical acceptability.

#### Evaluation approaches in text-to-text, image-to-text and evaluation methodology studies

Evaluations in text-to-text studies emphasise completeness, conciseness, and factuality, often combining ROUGE or BERTScore with medical expert evaluation.

In image-to-text settings, BLEU and BERTScore are frequently reported alongside domain-adapted measures such as CheXpert coverage and F1-RadGraph, with human review conducted on a subset of outputs. Regarding consistency across metrics, CIDEr-D and METEOR tends to track the lexical overlap metrics such as BLEU and ROUGE^37,38^, and F1-RadGraph follows the same overall trend as BLEU, ROUGE, and BERTScore ^39^.

Evaluation methodology studies place less weight on traditional lexical overlap metrics, favouring semantic and domain-adapted measures such as BERTScore, BARTScore, and the medical adaptations listed above, often supplemented by detailed expert assessments.

#### Language-Specific Limitations

Only 5 of 37 reviewed studies used non-English data (Supplementary Table S8). Three analysed Chinese, one Korean, and one Swedish text. Several papers did not state the language of the texts^40–42^. Zhang et al. (2023)^43^ explicitly considered language in their evaluation of tasks spanning English and Chinese; the faithfulness pipeline, FaR, relies on language-specific resources and procedures, for example UMLS for English and human tagging for Chinese, which illustrates the dependence of semantic factuality measures on the underlying language resources.

Lexical overlap metrics, such as BLEU and ROUGE, are sensitive to tokenisation and morphology. Character-level tokenisation, common in Chinese, produces different overlap values from word-level tokenisation in English. In morphologically rich languages, for example Finnish, inflection reduces n-gram overlap even when meaning is preserved, which limits comparability with English unless stemming or lemmatisation is applied. For embedding-based semantic metrics, performance is contingent on the underlying embedding model. In multilingual settings, for example the use of BGE in Zhang et al. (2025)^44^, the choice is appropriate to a Chinese context, although the checkpoint version was not reported. More generally, few studies specified the backbone or checkpoint, which constrains interpretability and limits cross-language comparability.

### Experimental Setup

#### Lexical Overlap Metrics

Lexical overlap metrics, including BLEU and ROUGE (1, 2, 4, and L), declined consistently as perturbation severity increased. This trend is shown in Figure 2 which displays BLEU scores (panels a-c) and ROUGE-L scores (panels d-f) across three perturbation levels for deletion, modification, and paraphrasing edits in English, Swedish, and Finnish. Even mild edits (level 1) reduced scores measurably, while severe edits (level 3) led to substantial drops across all metrics including paraphrasing that preserved meaning.

**Figure 2.**
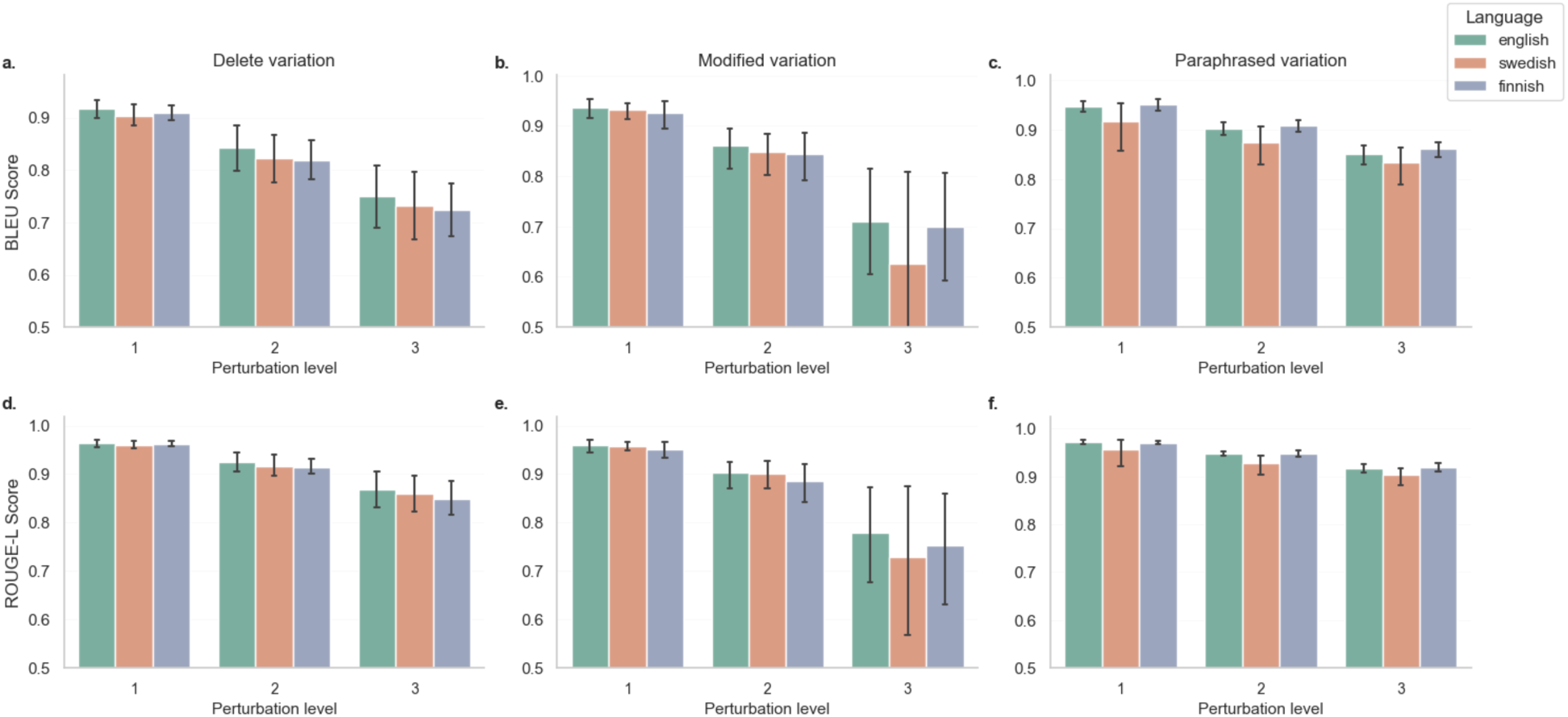
Lexical overlap metrics. Panels a-c: BLEU scores. Panels d–f: ROUGE-L scores. Bars show means with 95% confidence intervals across three perturbation levels for each model. Detailed breakdown of ROUGE-1, -2, -3, and -4 in Supplementary Figure S1.

BLEU was more sensitive to deletions and key-detail modifications than to paraphrase. Scores declined stepwise as deletion and modification levels increased, whereas paraphrase produced only modest reductions. ROUGE-L showed a similar pattern but was less sensitive overall, remaining relatively stable under paraphrase.

Higher-order overlap accentuated this contrast. ROUGE-4, which depends on longer n-grams, dropped sharply with extensive edits, especially deletions and sentence-level changes. ROUGE-1 remained comparatively stable, reflecting persistence of key vocabulary despite rewording.

#### Semantic Metrics

In addition to the most used semantic metrics BERTScore and BLEURT in our systematic review, we evaluated cosine similarity, given its foundational role among embedding-based measures. The results varied across perturbation types, severity levels, and languages (Figure 3). All metrics remained relatively high at perturbation level 1, particularly for modified and paraphrased variations, indicating that minimal edits preserved much of the original semantic content.

**Figure 3.**
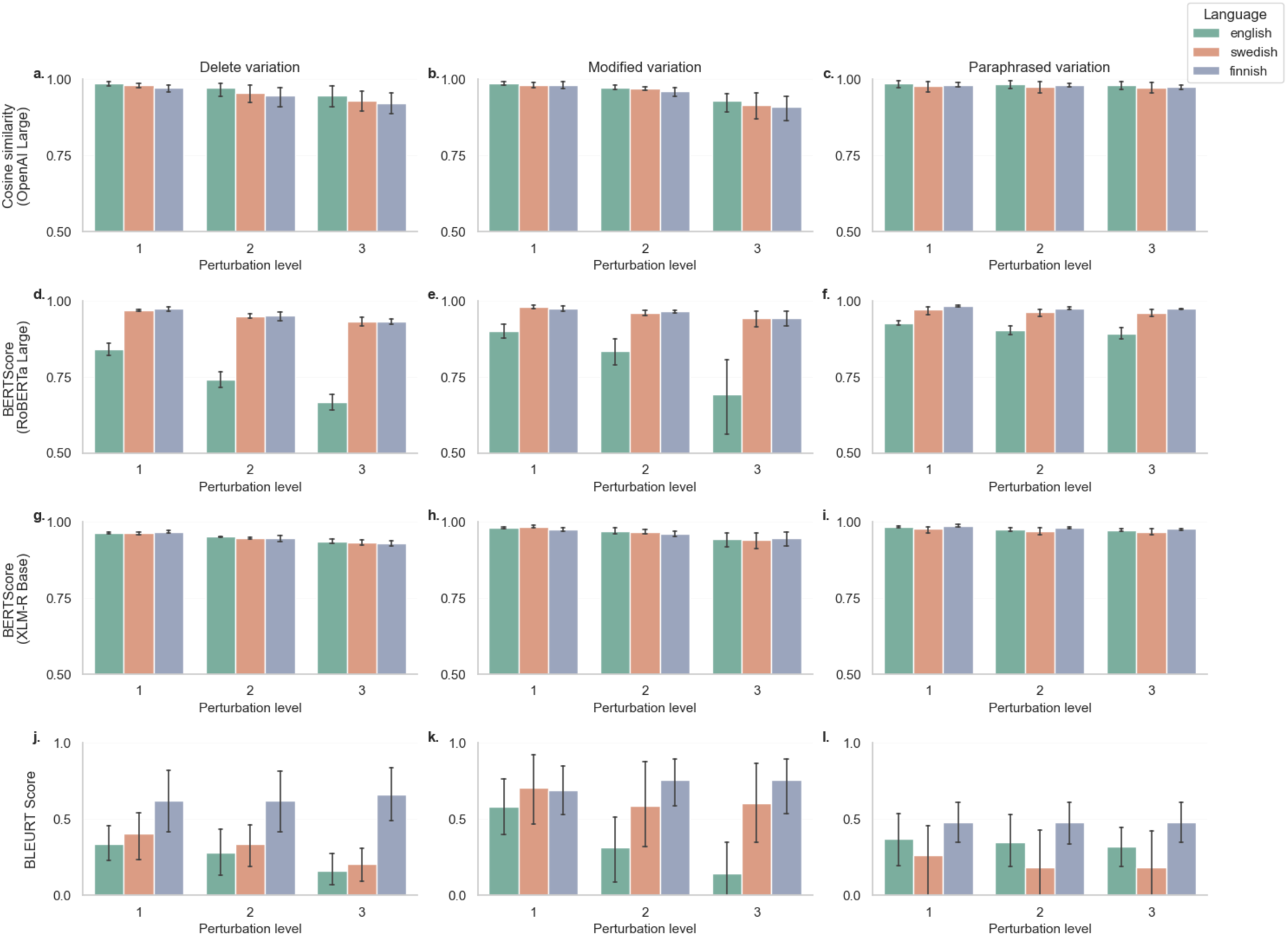
Semantic metrics by perturbation type and severity. Panels show mean BERTScore and BLEURT scores with 95% confidence intervals across three languages (English, Finnish, Swedish) for each perturbation type (deletion, modified, paraphrased) at increasing levels of perturbation (1 to 3). Results are grouped by semantic metric. Most showed decreasing similarity with higher perturbation levels, with limited differentiation between paraphrased and structurally altered inputs. Note: BLEURT score has a different y-axis.

As perturbation severity increased from levels 1 to 3, both BERTScore and BLEURT showed consistent declines across deletion, modification, and paraphrasing. Deletion-based perturbations tended to result in steeper drops, while paraphrased variants often retained higher scores.

Across languages, English and Swedish generally maintained higher semantic similarity scores compared to Finnish, which showed lower values across both metrics. Despite these differences, overall trends were consistent across the three languages, with increasing perturbation severity leading to decreased semantic alignment.

#### LLM-based Evaluation

LLM-as-evaluator change scores varied across models and languages in response to deletion and modification perturbations (Figure 4).

**Figure 4.**
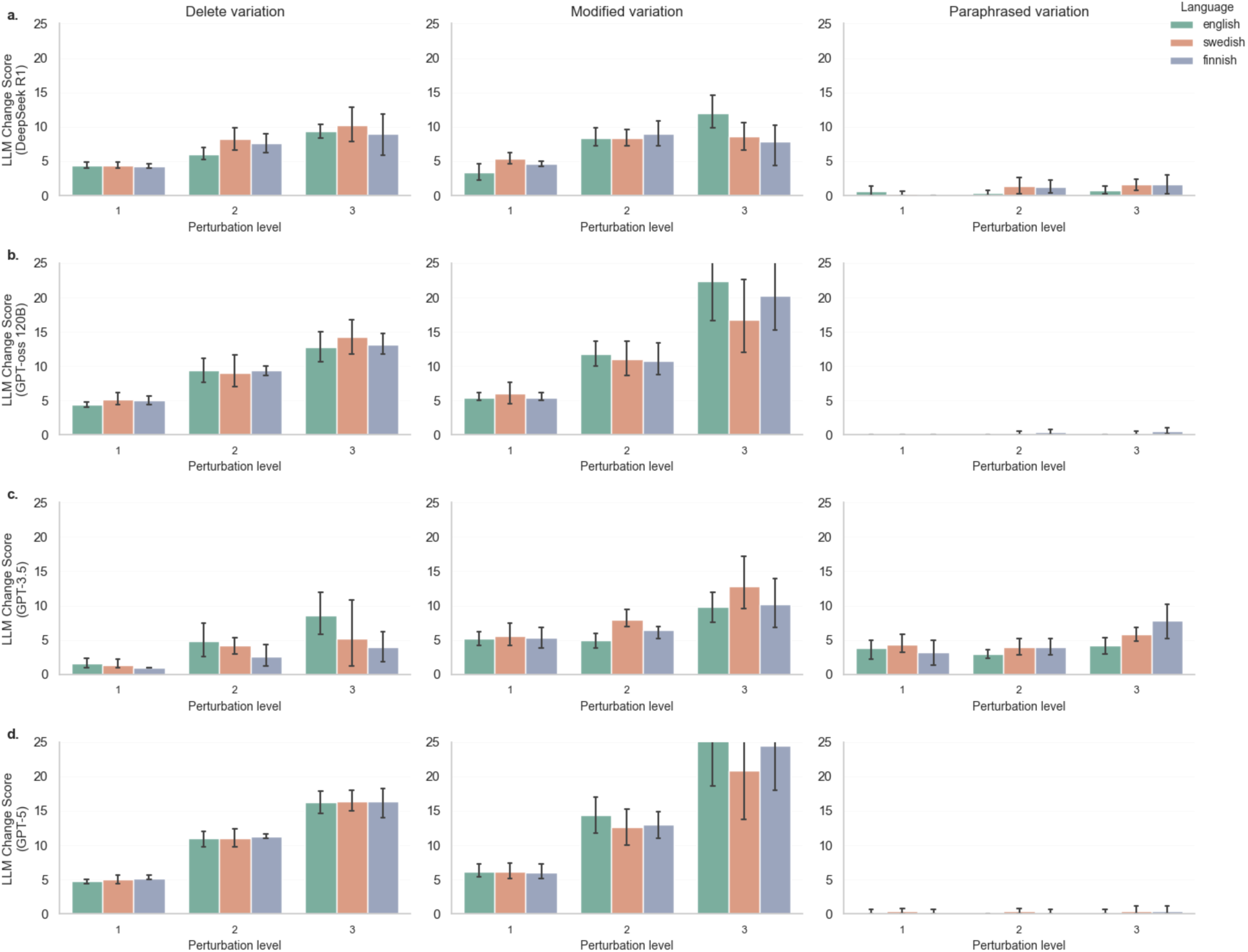
LLM-as-evaluator change scores for deletion and modification perturbations. Bars show means with 95% confidence intervals across three perturbation levels for each model. Models displayed in the main figure are GPT-5, GPT-4o, DeepSeek R1, gpt-oss-120B, and GPT-3.5. Additional models are provided in the supplement, including Gemma 3n E2B, Gemma 3n E4B, gpt-oss-20B, GPT-4, GPT-4.1, GPT-5 Mini and GPT-5 Nano (Supplementary Figure S2).

Paraphrased variants produced low change scores for most models. Two exceptions were Gemma3n E4B and GPT-3.5, which showed consistently elevated paraphrase scores across levels (Figure 4 and Supplementary Figure S2). The same pattern was observed for Gemma3n E2B.

All models showed monotonic increases in change scores with increasing deletion and modification levels. The magnitude of these increases was smaller for lower capacity models and larger for higher capacity models.

Variability across English, Swedish, and Finnish was greater for smaller models, with wider intervals and occasional language-specific deviations. Larger models yielded more consistent trajectories across languages with narrower intervals. English generally showed the tightest intervals.

Taken together, increasing model capacity was associated with improved paraphrase robustness, greater sensitivity to structural edits, and enhanced cross-language consistency.

#### Statistical Significance of the Experimental Results

Linear mixed-effects models, *metric~level* + (1|*case*), yielded clear and directionally consistent fixed effects for level (Table 2; full results in Supplementary Table S9).

**Table 2.** Fixed effect of perturbation level from linear mixed models. Points are β_level with 95% CI. ROUGE-L and BERTScore decrease with severity, GPT-5 increases. Full outputs in Supplementary Table S9.

| | ROUGE-L $\beta$<br>(95% CI) | ROUGE-L $p$-value | BERTScore $\beta$ (95% CI) | BERTScore $p$-value | GPT-5 $\beta$ (95% CI) | GPT-5 $p$-value |
| --- | --- | --- | --- | --- | --- | --- |
| English |  |  |  |  |  |  |
| Delete | -0.048<br>(-0.061 to -0.035) | <0.001 | -0.087<br>(-0.100 to -0.073) | <0.001 | 5.70<br>(4.99 to 6.41) | <0.001 |
| Modify | -0.090<br>(-0.131 to -0.049) | <0.001 | -0.104<br>(-0.155 to -0.053) | <0.001 | 10.00<br>(6.65 to 13.35) | <0.001 |
| Paraphrase | -0.027<br>(-0.031 to -0.024) | <0.001 | -0.017<br>(-0.023 to -0.011) | <0.001 | 0.00<br>(-0.24 to 0.24) | 1.00 |
| Swedish |  |  |  |  |  |  |
| Delete | -0.050<br>(-0.063 to -0.037) | <0.001 | -0.018<br>(-0.025 to -0.012) | <0.001 | 5.70<br>(5.18 to 6.22) | <0.001 |
| Modify | -0.115<br>(-0.179 to -0.051) | <0.001 | -0.019<br>(-0.028 to -0.009) | <0.001 | 7.30<br>(2.95 to 11.65) | 0.001 |
| Paraphrase | -0.027<br>(-0.032 to -0.021) | <0.001 | -0.005<br>(-0.008 to -0.002) | 0.001 | 0.00<br>(-0.24 to 0.24) | 1.00 |
| Finnish |  |  |  |  |  |  |
| Delete | -0.057<br>(-0.069 to -0.044) | <0.001 | -0.021<br>(-0.026 to -0.016) | <0.001 | 5.60<br>(4.62 to 6.58) | <0.001 |
| Modify | -0.098<br>(-0.143 to -0.054) | <0.001 | -0.015<br>(-0.025 to -0.006) | 0.002 | 9.20<br>(6.45 to 11.95) | <0.001 |
| Paraphrase | -0.025<br>(-0.029 to -0.022) | <0.001 | -0.005<br>(-0.007 to -0.003) | <0.001 | 0.10<br>(-0.06 to 0.26) | 0.21 |

#### Correlations

Normalized Spearman correlation coefficients (ρ) were calculated to assess how well each metric captured increasing perturbation severity across English, Swedish, and Finnish, separately for deletion, modification, and paraphrasing (Supplementary Figure S3).

For modified and deleted summaries (Supplementary Figure S3a and b), most metrics demonstrated strong positive correlations with perturbation level across all three languages. BLEU and ROUGE-L consistently showed high responsiveness to structural edits (ρ = 0·81-0·87). Cosine similarity had a noticeable difference between modifications and deletions, were modifications led to higher scores. BERTScore tracked perturbations reliably, although Finnish modifications produced noticeably weaker correlations. BLEURT performed poorly, yielding consistently low values and even negative correlations in Finnish, indicating a failure to register structural differences. English modifications generally showed the strongest correlations, whereas deletions did not reveal such a language-specific pattern.

LLM-as-evaluator metrics varied across models. GPT-4 and GPT-5, along with their variants, were the most consistent, reaching ρ values up to 0·96 across languages. In contrast, GPT-3.5 showed reduced performance for Swedish deletions (ρ = 0·35). DeepSeek R1 performed moderately, with strong correlations for English deletions and modifications (ρ = 0·88-0·89), but substantially lower values for Swedish and especially Finnish inputs.

For paraphrased summaries (Supplementary Figure S3c), correlations were considerably weaker. BLEU and ROUGE-L reached high ρ values (up to 0·94), but these inflated scores reflected their sensitivity to surface-level differences rather than true semantic similarity. Semantic metrics showed reduced reactivity to paraphrasing, particularly in Swedish (ρ = 0·25-0·49), though some correlation was retained. Cosine similarity was the semantic metric with the lowest reactivity to paraphrasing and therefore performed best in this aspect. BERTScore was moderately sensitive to paraphrasing across languages, but consistently lower than for deletions or modifications. BLEURT, in contrast, was largely unresponsive, showing minimal correlation across all languages. While this lack of reactivity could be desirable for paraphrases, BLEURT also failed to capture structural edits, limiting its utility. GPT-4 and GPT-4o appropriately showed minimal or no correlation for paraphrased content, suggesting robustness to lexical variation. By contrast, GPT-3·5 and DeepSeek R1 produced inconsistent results, with moderate to high correlations in Swedish and Finnish but little sensitivity in English, reflecting undesired variability.

## Discussion

This review shows a field in transition. The most commonly used lexical overlap metrics, chiefly ROUGE and BLEU, do not reliably reflect clinical meaning in this setting, as demonstrated by our experimental setup. They are attractive because they are simple to compute, low in computational cost, widely familiar, and facilitate comparisons across datasets, yet in our experimental setup they penalised meaning-preserving paraphrasing and therefore mis-ranked outputs that retained the same clinical content. In contrast, semantic metrics and LLM-based evaluators were more robust to rewording and tracked clinically relevant changes, with absolute sensitivity varying by model. LLM-based evaluations using large models such as DeepSeek-R1, GPT-oss-120B, and GPT-5, in particular, penalised deletions and modifications of key clinical details while applying little or no penalty to paraphrase, aligning more closely with safety-critical needs.

Language effects were pronounced. Most evidence comes from English, which limits generalisability to low-resource languages. Tokenisation, morphology, and word order differ across languages and can alter metric behaviour. In our data, Finnish showed lower absolute scores and weaker monotonic trends, consistent with agglutinative morphology and variable syntax, whereas Swedish behaved more like English. Evaluators, whether lexical overlap, embedding-based or LLM-based, should therefore be selected and validated within each language.

Reporting practices remain inconsistent. Several articles cited ‘ROUGE’ or ‘BLEU’ without specifying the variant, and BERTScore results were sometimes reported without the embedding model or adequate model details. Such omissions hinder reproducibility and comparability. We recommend reporting the exact metric variant and configuration, and the embedding model and checkpoint.

A practical constraint is computational cost. In principle, a small, low-resource model that performs well would be preferable, but in practice performance often improves with capacity. We observed the same pattern: smaller encoders and lower-capacity LLMs were more sensitive to paraphrasing and less sensitive to deletions and key modifications, an undesirable profile for clinical evaluation. Training data also matters. A compact model trained on the target language can, in some settings, outperform a larger model trained predominantly on English. In our BERTScore experiments, however, RoBERTa-large preserved semantic similarity under paraphrase more effectively than XLM-R base across all languages, despite the latter’s multilingual training.

We favour LLM-based evaluators as the primary judge because they track clinically relevant changes while remaining tolerant of paraphrase. When resources are constrained, semantic metrics can provide a rough, low-overhead first screen for meaning preservation, after which a narrowed subset can be escalated to an LLM evaluator. Because evaluation is language dependent, both semantic metrics and the LLM-as-evaluators should be selected and validated within each language. Absolute cross-language score comparisons should be avoided in favour of within-language contrasts. Sensitivity analyses across institutions, specialties, time periods, and patient subgroups will strengthen claims of generalisability.

Finally, although LLM-based evaluation remains uncommon and has largely been applied to simpler tasks, results from our experimental setup indicate that it can automate domain-specific evaluation when carefully specified and constrained. Wider adoption will require prospective validation against clinician judgement, explicit controls for length and formatting effects, and systematic bias assessment and mitigation. Overall, the evidence supports a shift from lexical overlap metrics to meaning-aware, domain-adapted evaluation, preferably LLM-based where feasible, with rigorous reporting and language-sensitive design.

## Limitations

The evidence base was heterogeneous in design, setting, and outcome definitions, and reporting was often incomplete, which precluded meta-analysis and limits cross-study comparability. Many primary studies used small samples, lacked blinded assessment, or did not report inter-rater reliability, reducing confidence in observed effects. Few papers evaluated image-to-text generation or focused explicitly on evaluation methodology. Several studies reported only generic metric labels, for example “ROUGE” without a variant, or omitted the embedding backbone, which constrains reproducibility.

We searched two databases to 10 April 2025 and restricted inclusion to peer-reviewed English-language publications, so publication and language bias are possible. Although two reviewers screened with a third checking all decisions, harmonising heterogeneous terminology required judgement and may have introduced misclassification despite predefined rules. When information was missing or unclear, we recorded it as not reported and documented any assumptions made for synthesis, which we flag in the Results and Supplement. We used narrative synthesis because effect measures were not comparable, and we did not apply formal outcome-level risk-of-bias or certainty grading. Newer studies may have appeared after our search window.

Two issues complicate the synthesis of the findings of the reviewed papers. Foremost, terminology across studies was inconsistent and sometimes insufficiently defined: terms “faithfulness,” “accuracy,” and “factuality” were used to refer to a variety of concepts, respectively, including sometimes interchangeably, and “semantics” referred to linguistic meaning, computational entailment, or concept coverage. Additionally, when criteria were not clearly defined, the rationale connecting them to evaluation approaches was sometimes unclear, particularly in studies employing several criteria and approaches. Thereafter, few studies prioritized the relative importance of different types of errors, for example distinguishing clinically critical omissions from minor stylistic deviations.

The experimental setup used synthetic clinical cases and controlled perturbations that capture deletions and substitutions, but they may not reflect pragmatic failure modes such as temporal reasoning errors, guideline adherence, or local documentation conventions. The sample was limited to five internal medicine cases in three languages, which restricts generalisability to other note types, specialties, and languages. Metric behaviour depends on implementation choices, for example tokenisation, smoothing, and checkpoint selection, which, although versioned in the repository, may differ from other groups’ setups. LLM-as-evaluator results can be sensitive to prompts, decoding limits, and API version drift; we mitigated this with fixed prompts and documented settings, yet residual variability remains. The experiment evaluates text quality signals rather than clinical outcomes, workflow impact, or patient harm, so it should be interpreted as a methodological probe rather than a test of clinical effectiveness or acceptability.

## Conclusions

Evaluation of AI-generated clinical documentation is moving from lexical overlap metrics to metrics that capture meaning and clinical relevance yet practice still trails clinical requirement. Evaluations should explicitly detect and quantify erroneous perturbations, for example hallucinated additions, altered clinical facts, and clinically relevant omissions, since these may carry significant clinical consequences. When compute is limited, a practical battery prioritises semantic, embedding-based metrics as the initial screen for meaning preservation; when feasible, prefer an LLM-as-evaluator for the primary assessment. Given the speed and volume of AI-generated text, reserve structured human review for critical samples and use LLM-as-evaluator for scale with agreement statistics reported and periodic revalidation. To improve reproducibility, studies should specify the exact metric configurations, embedding models and checkpoints. Given the predominance of English-language research relative to care delivered in local languages, there is a particular need for evaluations conducted in non-English settings.

## Data Availability

All data generated in this study are available within the published article or its supplementary materials. The source code supporting this study is accessible at https://github.com/achdahlberg/quality-AI-gen-clinical-notes-2025.

## Contributions

AD, TK, and TW-J conducted all stages of the systematic review described in Methods, performed screening and synthesis, and drafted the review results. AD led conceptualisation and study design, prepared the Finnish discharge summaries and generated the dataset, developed and executed most components of the experimental setup, and wrote the experimental Methods and Results. RL contributed to the conceptualisation of the experimental framework, ran most closed-source LLM evaluations, and critically revised the manuscript. OT and TP assisted with the experimental work, provided methodological guidance and code implementation, and critically revised the manuscript. MG designed, coordinated and coded the MedBench data base used in the study, and reviewed the manuscript. VV and ES provided strategic input on study design and methodology, supervised the research, contributed to manuscript development, and critically revised the manuscript. All authors contributed to interpretation of the findings, approved the final version, and agree to be accountable for the work.

## Acknowledgements

The authors gratefully acknowledge the creators of the original discharge summaries included in the MedBench dataset. This work was supported by Business Finland (1562/31/2024) through the GenAID research project. AD acknowledges Frans Wilhelm och Waldermar von Frenckells understödsfond and The Finnish Medical Foundation for the award of a personal grant. ES acknowledges Finska Läkaresällskapet, The Finnish Medical Foundation and, Liv och Hälsa Medical Support Association.

## Declaration of generative AI and AI-assisted technologies in the manuscript preparation process

During the preparation of this work, the authors used ChatGPT (OpenAI)^45^ and Cursor AI Coding Assistant^46^. These tools were employed to improve the clarity and coherence of the language, and work with some routine parts of the required programming tasks. The assistants were only involved in base code editing work and were not used for any insights or exploration. After using these tools, the authors reviewed and edited the content as needed and take full responsibility for the content of the published article.

## Supplementary Information

**Supplementary Figure S1.**
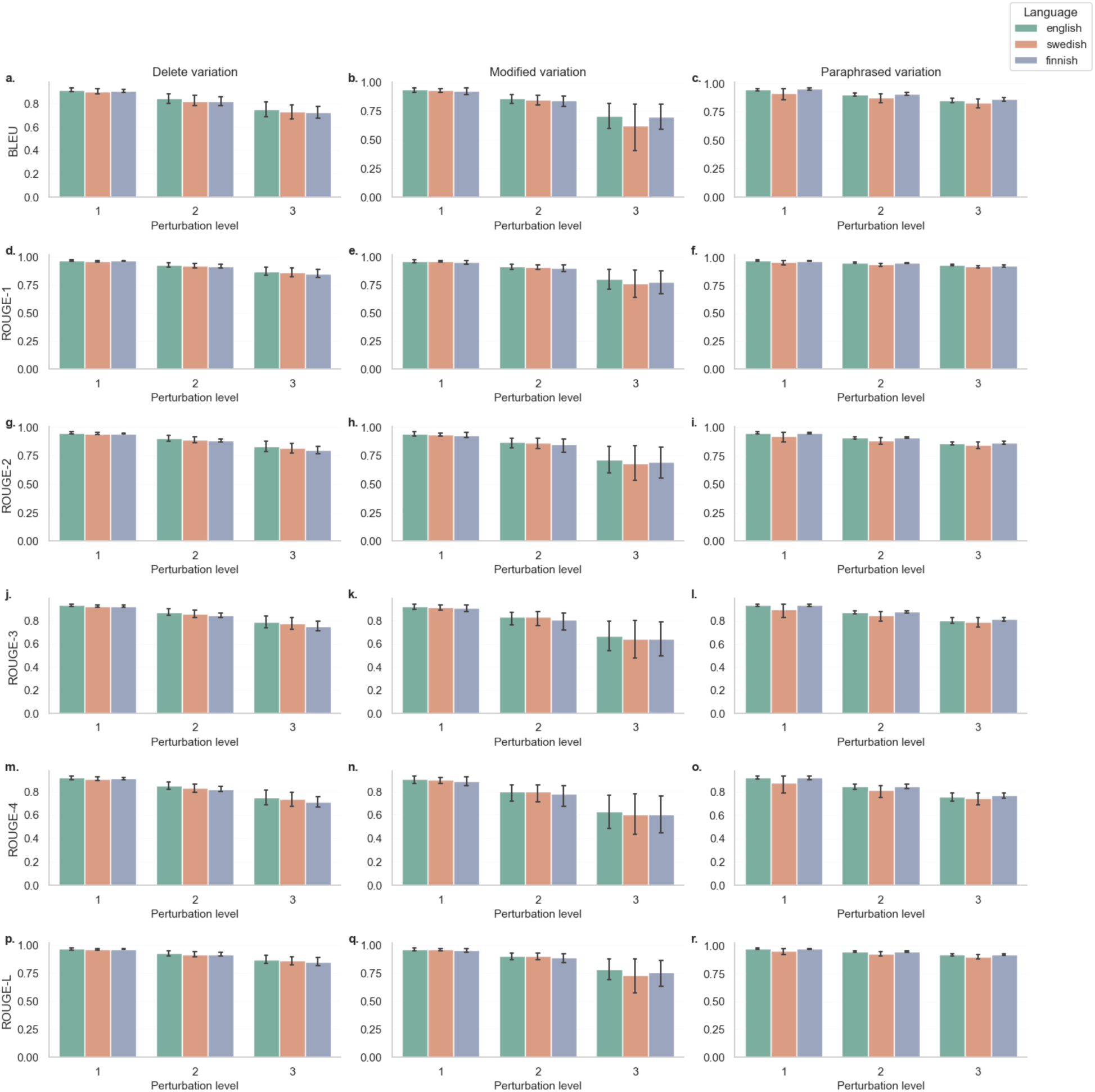
Lexical overlap metrics.

**Supplementary Figure S2.**
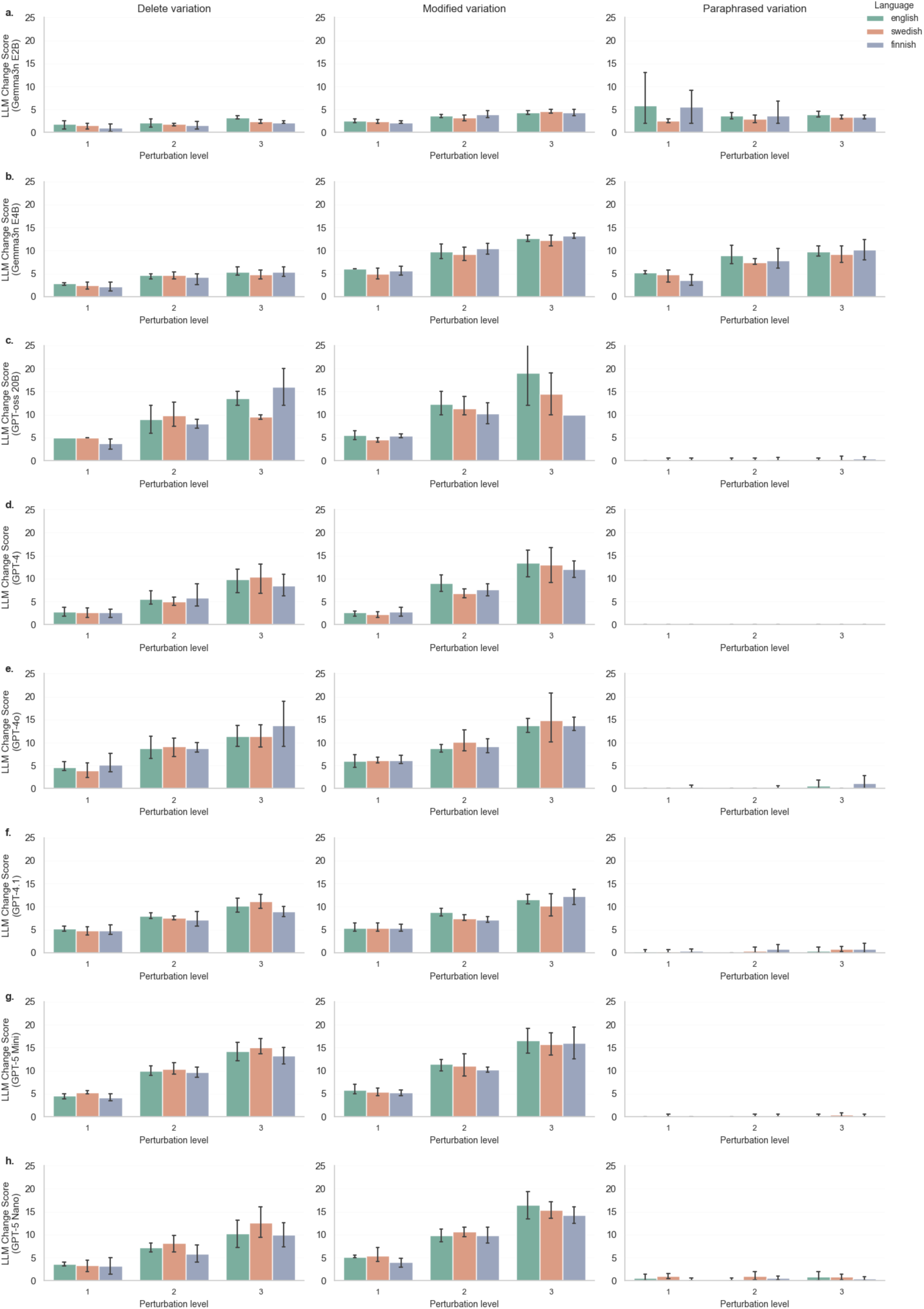
LLM-as-evaluator.

**Supplementary Figure S3.**
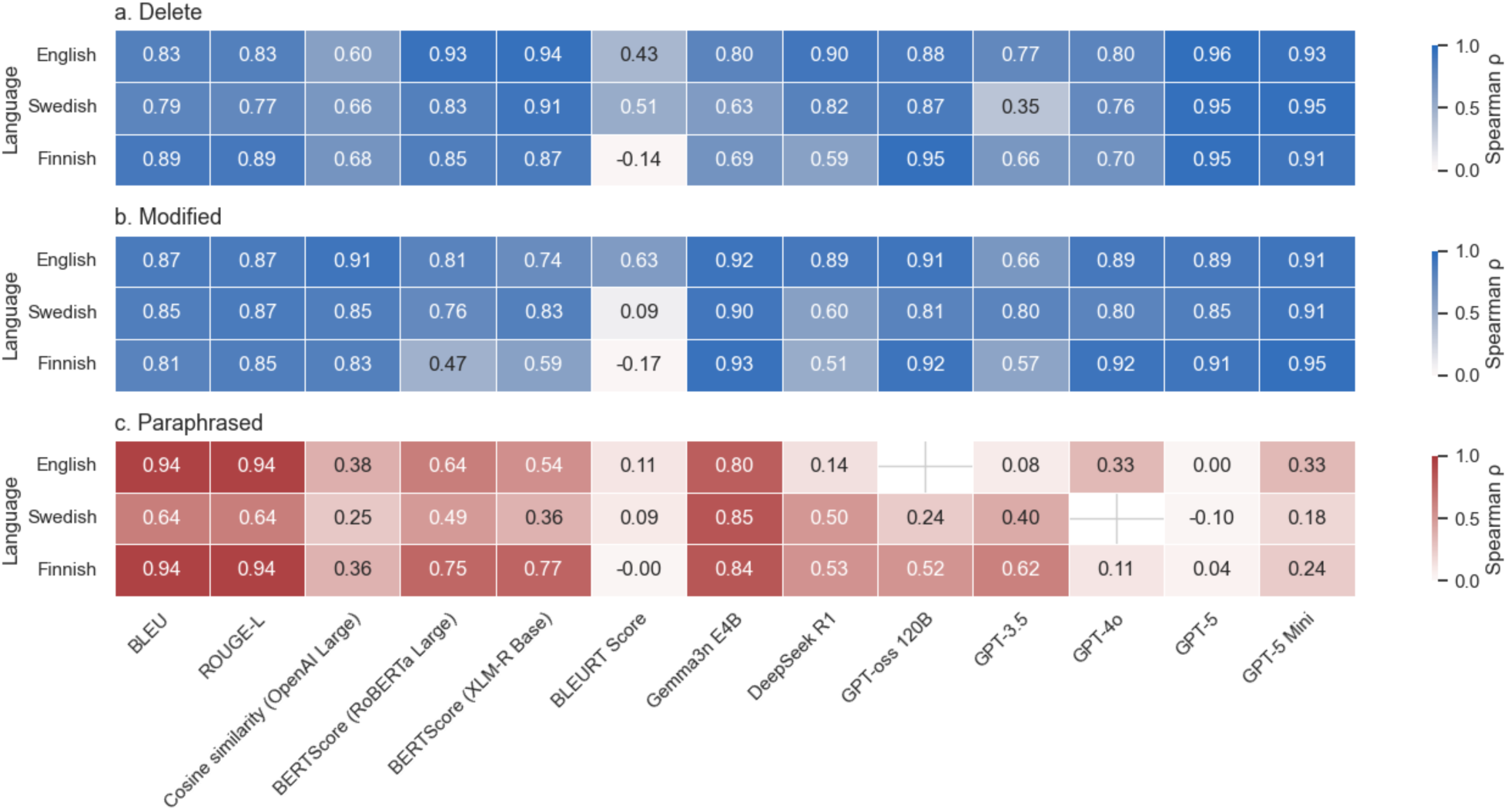
Spearman P.

**Supplementary Table S1.** PRISMA checklist^1^.

| Section and Topic | Item # | Checklist item | Location where item is reported |
| --- | --- | --- | --- |
| **TITLE** |  |  |  |
| Title | 1 | Identify the report as a systematic review. | 1 |
| **ABSTRACT** |  |  |  |
| Abstract | 2 | See the PRISMA 2020 for Abstracts checklist. | 1-2 |
| **INTRODUCTION** |  |  |  |
| Rationale | 3 | Describe the rationale for the review in the context of existing knowledge. | 2-4 |
| Objectives | 4 | Provide an explicit statement of the objective(s) or question(s) the review addresses. | 4 |
| **METHODS** |  |  |  |
| Eligibility criteria | 5 | Specify the inclusion and exclusion criteria for the review and how studies were grouped for the syntheses. | 5-6 |
| Information sources | 6 | Specify all databases, registers, websites, organisations, reference lists and other sources searched or consulted to identify studies. Specify the date when each source was last searched or consulted. | 4-5 |
| Search strategy | 7 | Present the full search strategies for all databases, registers and websites, including any filters and limits used. | 4-5, supplements |
| Selection process | 8 | Specify the methods used to decide whether a study met the inclusion criteria of the review, including how many reviewers screened each record and each report retrieved, whether they worked independently, and if applicable, details of automation tools used in the process. | 6 |
| Data collection process | 9 | Specify the methods used to collect data from reports, including how many reviewers collected data from each report, whether they worked independently, any processes for obtaining or confirming data from study investigators, and if applicable, details of automation tools used in the process. | 7-8 |
| Data items | 10a | List and define all outcomes for which data were sought. Specify whether all results that were compatible with each outcome domain in each study were sought (e.g. for all measures, time points, analyses), and if not, the methods used to decide which results to collect. | 7-8 |
|  | 10b | List and define all other variables for which data were sought (e.g. participant and intervention characteristics, funding sources). Describe any assumptions made about any missing or unclear information. | 7-8 |
| Study risk of bias assessment | 11 | Specify the methods used to assess risk of bias in the included studies, including details of the tool(s) used, how many reviewers assessed each study and whether they worked independently, and if applicable, details of automation tools used in the process. | 7-8 |
| Effect measures | 12 | Specify for each outcome the effect measure(s) (e.g. risk ratio, mean difference) used in the synthesis or presentation of results. | 8-9 |
| Synthesis methods | 13a | Describe the processes used to decide which studies were eligible for each synthesis (e.g. tabulating the study intervention characteristics and comparing against the planned groups for each synthesis (item #5)). | 8-9 |
|  | 13b | Describe any methods required to prepare the data for presentation or synthesis, such as handling of missing summary statistics, or data conversions. | 8-9 |
|  | 13c | Describe any methods used to tabulate or visually display results of individual studies and syntheses. | 8-9 |
|  | 13d | Describe any methods used to synthesize results and provide a rationale for the choice(s). If meta-analysis was performed, describe the model(s), method(s) to identify the presence and extent of statistical heterogeneity, and software package(s) used. | 8-9 |
|  | 13e | Describe any methods used to explore possible causes of heterogeneity among study results (e.g. subgroup analysis, meta-regression). | 8-9 |
|  | 13f | Describe any sensitivity analyses conducted to assess robustness of the synthesized results. | 8-9 |
| Reporting bias assessment | 14 | Describe any methods used to assess risk of bias due to missing results in a synthesis (arising from reporting biases). | 8-9 |
| Certainty assessment | 15 | Describe any methods used to assess certainty (or confidence) in the body of evidence for an outcome. | 8-9 |
| **RESULTS** |  |  |  |
| Study selection | 16a | Describe the results of the search and selection process, from the number of records identified in the search to the number of studies included in the review, ideally using a flow diagram. | 6-7 |
|  | 16b | Cite studies that might appear to meet the inclusion criteria, but which were excluded, and explain why they were excluded. | 7 |
| Study characteristics | 17 | Cite each included study and present its characteristics. | Supplementary Table S3 |
| Risk of bias in studies | 18 | Present assessments of risk of bias for each included study. | Not included, see p. 4 |
| Results of individual studies | 19 | For all outcomes, present, for each study: (a) summary statistics for each group (where appropriate) and (b) an effect estimate and its precision (e.g. confidence/credible interval), ideally using structured tables or plots. | Supplementary Table S3 |
| Results of syntheses | 20a | For each synthesis, briefly summarise the characteristics and risk of bias among contributing studies. | Supplementary Table S3 |
|  | 20b | Present results of all statistical syntheses conducted. If meta-analysis was done, present for each the summary estimate and its precision (e.g. confidence/credible interval) and measures of statistical heterogeneity. If comparing groups, describe the direction of the effect. | - |
|  | 20c | Present results of all investigations of possible causes of heterogeneity among study results. | 18 |
|  | 20d | Present results of all sensitivity analyses conducted to assess the robustness of the synthesized results. | - |
| Reporting biases | 21 | Present assessments of risk of bias due to missing results (arising from reporting biases) for each synthesis assessed. | - |
| Certainty of evidence | 22 | Present assessments of certainty (or confidence) in the body of evidence for each outcome assessed. | - |
| **DISCUSSION** |  |  |  |
| Discussion | 23a | Provide a general interpretation of the results in the context of other evidence. | 31-32 |
|  | 23b | Discuss any limitations of the evidence included in the review. | 33 |
|  | 23c | Discuss any limitations of the review processes used. | 33-34 |
|  | 23d | Discuss implications of the results for practice, policy, and future research. |  |
| **OTHER INFORMATION** |  |  |  |
| Registration and protocol | 24a | Provide registration information for the review, including register name and registration number, or state that the review was not registered. | 4 |
|  | 24b | Indicate where the review protocol can be accessed, or state that a protocol was not prepared. | 4 |
|  | 24c | Describe and explain any amendments to information provided at registration or in the protocol. | - |
| Support | 25 | Describe sources of financial or non-financial support for the review, and the role of the funders or sponsors in the review. | 34 |
| Competing interests | 26 | Declare any competing interests of review authors. | 35 |
| Availability of data, code and other materials | 27 | Report which of the following are publicly available and where they can be found: template data collection forms; data extracted from included studies; data used for all analyses; analytic code; any other materials used in the review. | 34 |

**Supplementary Table S2.**
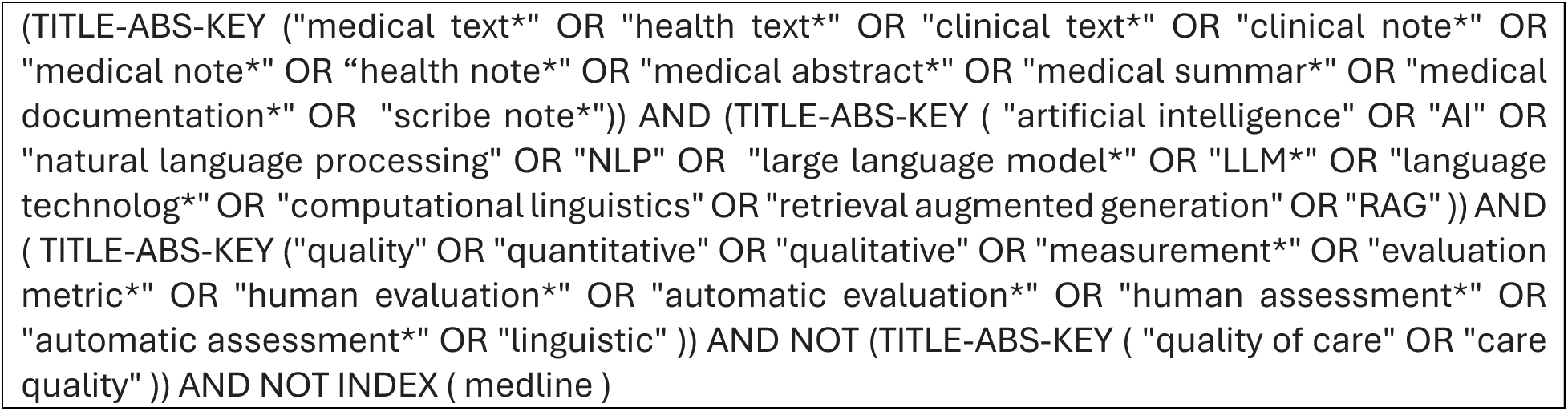
Search terms in Ovid MEDLINE and Scopus.

**Supplementary Table S3.** Eligibility criteria.

| Inclusion criteria | Exclusion criteria |
| --- | --- |
| - Language of the paper (English)<br>- Peer reviewed<br>- Any medical profession and specialty<br>- Any type of clinical note or transcription<br>- All care settings (hospital, pre-hospital, primary care)<br>- Original research articles or conference papers<br>- Includes evaluation of resulting clinical note | - Not focused on clinical documentation<br>- Focus on research, abstracts or other non-clinical texts<br>- Reviews, contextual papers, editorials, letters<br>- Audits<br>- Translation or simplification tasks<br>- Texts targeted at laypeople, not medical professionals<br>- Use of structured data to assess quality<br>- Quality measured only via outcomes or output volume<br>- Studies on documentation processes, systems, or training<br>- Duplicate publications |

**Supplementary Table S4.**
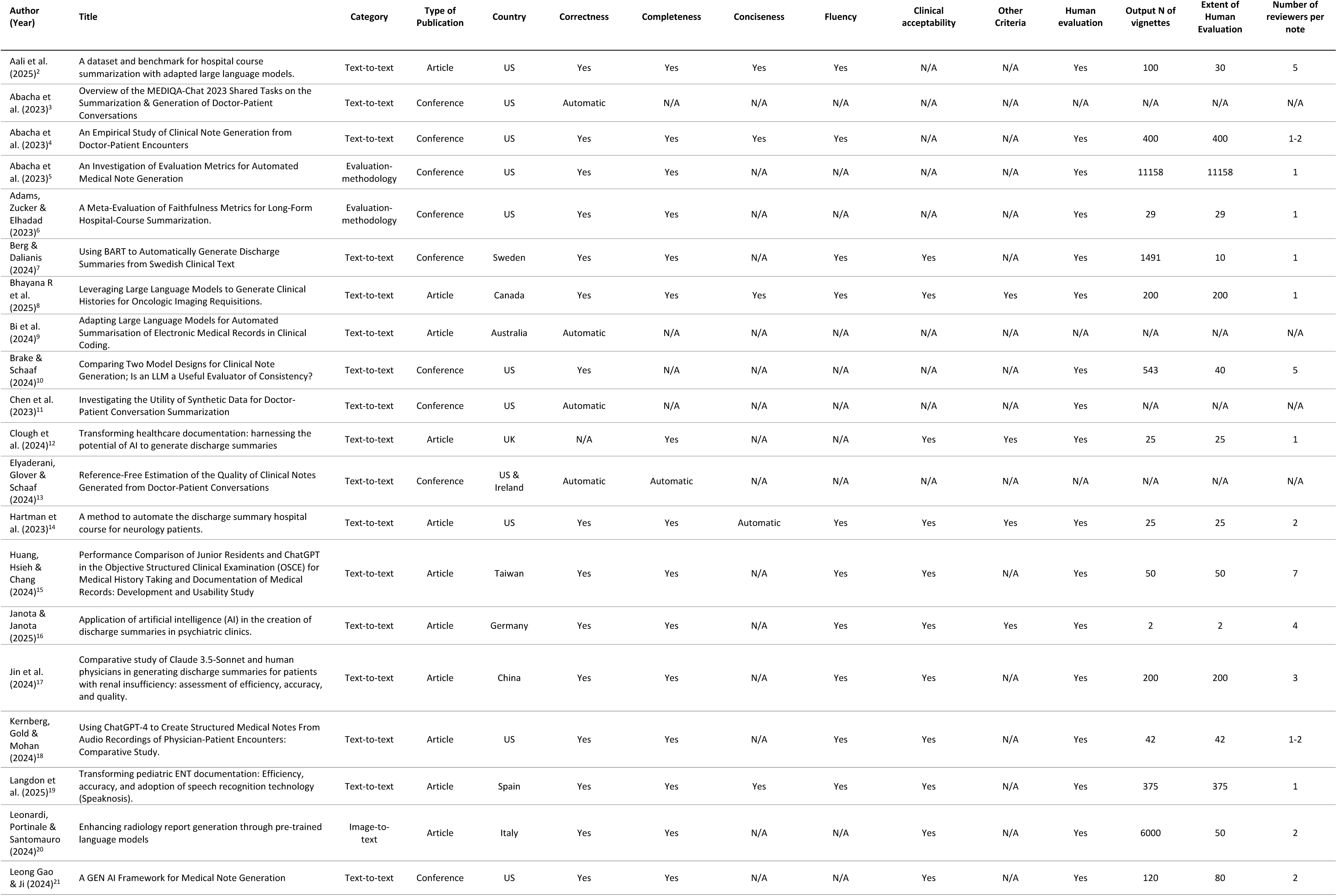
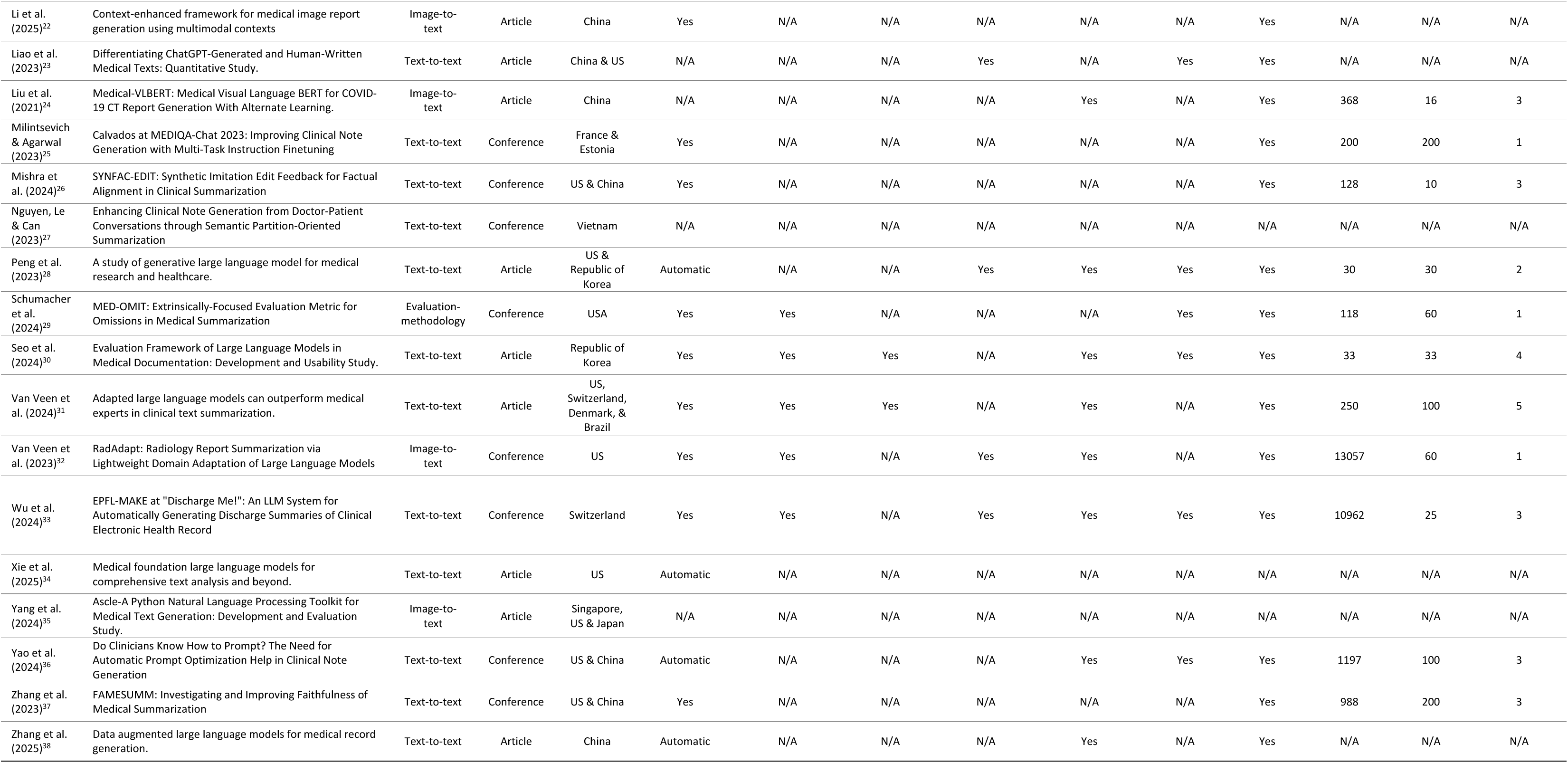
All included studies.

**Supplementary Table S5.** Automated metrics (lexical overlap and semantic metrics), which studies.

| Automated Metric | Studies |
| --- | --- |
| BLEU-1 | Li, Wang <sup>22</sup>, & Liu, Liao <sup>24</sup> |
| BLEU-2 | Li, Wang <sup>22</sup>, & Liu, Liao <sup>24</sup> |
| BLEU-3 | Li, Wang <sup>22</sup>, & Liu, Liao <sup>24</sup> |
| BLEU-4 | Leonardi, Portinale <sup>20</sup>, Li, Wang <sup>22</sup>, Liu, Liao <sup>24</sup>, & Wu, Boulenger <sup>33</sup> |
| BLEU | Aali, Van Veen <sup>2</sup>, Van Veen, Van Uden <sup>39</sup>, & Van Veen, Van Uden <sup>31</sup> |
| ROUGE-1 | Abacha, Yim <sup>3</sup>, Abacha, Fan <sup>4</sup>, Berg and Dalianis <sup>7</sup>, Bi, Liu <sup>9</sup>, Brake and Schaaf <sup>10</sup>, Chen, Grambow <sup>11</sup>, Hartman, Bapat <sup>14</sup>, Leong, Gao <sup>21</sup>, Milintsevich and Agarwal <sup>25</sup>, Mishra, Yao <sup>26</sup>, Nguyen, Le <sup>27</sup>, Wu, Boulenger <sup>33</sup>, Yang, Zeng <sup>35</sup>, Yao, Jaafar <sup>36</sup>, Zhang, Zhang <sup>37</sup>, & Zhang, Zhao <sup>38</sup> |
| ROUGE-2 | Abacha, Yim <sup>3</sup>, Abacha, Fan <sup>4</sup>, Berg and Dalianis <sup>7</sup>, Bi, Liu <sup>9</sup>, Brake and Schaaf <sup>10</sup>, Chen, Grambow <sup>11</sup>, Hartman, Bapat <sup>14</sup>, Leong, Gao <sup>21</sup>, Milintsevich and Agarwal <sup>25</sup>, Mishra, Yao <sup>26</sup>, Nguyen, Le <sup>27</sup>, Yang, Zeng <sup>35</sup>, Yao, Jaafar <sup>36</sup>, Zhang, Zhang <sup>37</sup>, & Zhang, Zhao <sup>38</sup> |
| ROUGE-3 | Brake and Schaaf <sup>10</sup>, Chen, Grambow <sup>11</sup> |
| ROUGE-4 | Chen, Grambow <sup>11</sup> |
| ROUGE-L | Aali, Van Veen <sup>2</sup>, Abacha, Yim <sup>3</sup>, Abacha, Fan <sup>4</sup>, Berg and Dalianis <sup>7</sup>, Bi, Liu <sup>9</sup>, Brake and Schaaf <sup>10</sup>, Chen, Grambow <sup>11</sup>, Hartman, Bapat <sup>14</sup>, Leonardi, Portinale <sup>20</sup>, Leong, Gao <sup>21</sup>, Li, Wang <sup>22</sup>, Liu, Liao <sup>24</sup>, Milintsevich and Agarwal <sup>25</sup>, Mishra, Yao <sup>26</sup>, Nguyen, Le <sup>27</sup>, Van Veen, Van Uden <sup>32</sup>, Van Veen, Van Uden <sup>31</sup>, Wu, Boulenger <sup>33</sup>, Xie, Chen <sup>34</sup>, Yang, Zeng <sup>35</sup>, Yao, Jaafar <sup>36</sup>, Zhang, Zhang <sup>37</sup>, & Zhang, Zhao <sup>38</sup> |
| ROUGE-Lsum | Leong, Gao <sup>21</sup>, Mishra, Yao <sup>26</sup>, Nguyen, Le <sup>27</sup>, Schumacher, Rosenthal <sup>29</sup> |
| BERTScore | Aali et al. (2025), Abacha et al. (2023), Adams, Zucker & Elhadad et al. (2023), Ben Abacha et al. (2023), Bi et al. (2024), Chen et al. (2023), Langdon et al. (2025), Leonardi, Portinale & Santomauro et al. (2024), Leong Gao & Ji et al. (2024), Milintsevich & Agarwal et al. (2023), Schumacher et al. (2024), Van Veen et al. (2023), Van Veen et al. (2024), Wu et al. (2024), Xie et al. (2025), Zhang et al. (2023) |
| BLEURT | Abacha et al. (2023), Ben Abacha et al. (2023), Leong Gao & Ji et al. (2024), Milintsevich & Agarwal et al. (2023) |
| BARTScore | Abacha et al. (2023), Adams, Zucker & Elhadad et al. (2023) |

**Supplementary Table S6.** Human evaluation extent. The first row summarises all vignettes evaluated by either automated or human methods. All subsequent rows refer only to studies that included human evaluation.

|  | Text-to-text | Image-to-text | Evaluation methodology |
| --- | --- | --- | --- |
| Total vignettes evaluated (both automatic and human), median (IQR) | 200 (42-400) | 6000 (3184–9528) | 118 (74–5638) |
| N of vignettes with human evaluation, median (IQR) | 42 (25–200) | 50 (33-55) | 60 (44-5609) |
| Median number of reviewers per vignette (IQR) | 3 (1·5-4·0) | 2 (1·5-2·5) | 1 (1·0-1·0) |
| Studies with >50% vignettes human-reviewed (% of human reviewed studies) | 13 (62%) | 0 (0%) | 3 (100%) |
| Studies with <50% vignettes human-reviewed (% of human reviewed studies) | 8 (38%) | 3 (100%) | 0 (0%) |
| **Human evaluation of all studies (% of n<sub>x</sub>)** | **21/29 (72%)** | **3/5 (60%)** | **3/3 (100%)** |

**Supplementary Table S7.**
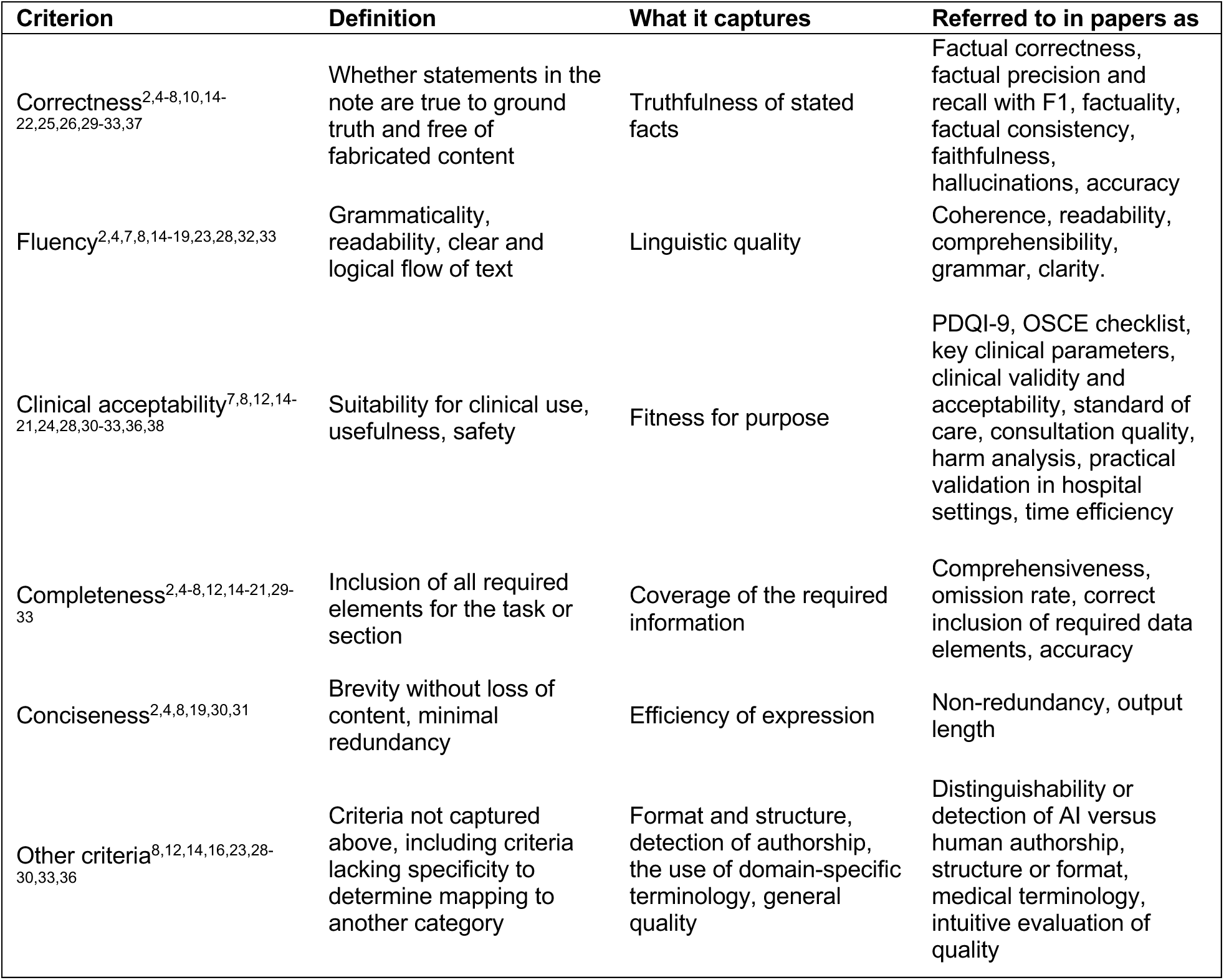
Evaluation criteria, preferred terms and definitions. The diverse labels used in the studies were mapped to the preferred criteria above. When a study used “accuracy” ambiguously, we assigned it to factual correctness or completeness based on context.

| Criterion | Definition | What it captures | Referred to in papers as |
| --- | --- | --- | --- |
| Correctness<sup>2,4-8,10,14-22,25,26,29-33,37</sup> | Whether statements in the note are true to ground truth and free of fabricated content | Truthfulness of stated facts | Factual correctness, factual precision and recall with F1, factuality, factual consistency, faithfulness, hallucinations, accuracy |
| Fluency<sup>2,4,7,8,14-19,23,28,32,33</sup> | Grammaticality, readability, clear and logical flow of text | Linguistic quality | Coherence, readability, comprehensibility, grammar, clarity. |
| Clinical acceptability<sup>7,8,12,14-21,24,28,30-33,36,38</sup> | Suitability for clinical use, usefulness, safety | Fitness for purpose | PDQI-9, OSCE checklist, key clinical parameters, clinical validity and acceptability, standard of care, consultation quality, harm analysis, practical validation in hospital settings, time efficiency |
| Completeness<sup>2,4-8,12,14-21,29-33</sup> | Inclusion of all required elements for the task or section | Coverage of the required information | Comprehensiveness, omission rate, correct inclusion of required data elements, accuracy |
| Conciseness<sup>2,4,8,19,30,31</sup> | Brevity without loss of content, minimal redundancy | Efficiency of expression | Non-redundancy, output length |
| Other criteria<sup>8,12,14,16,23,28-30,33,36</sup> | Criteria not captured above, including criteria lacking specificity to determine mapping to another category | Format and structure, detection of authorship, the use of domain-specific terminology, general quality | Distinguishability or detection of AI versus human authorship, structure or format, medical terminology, intuitive evaluation of quality |

**Supplementary Table S8.**
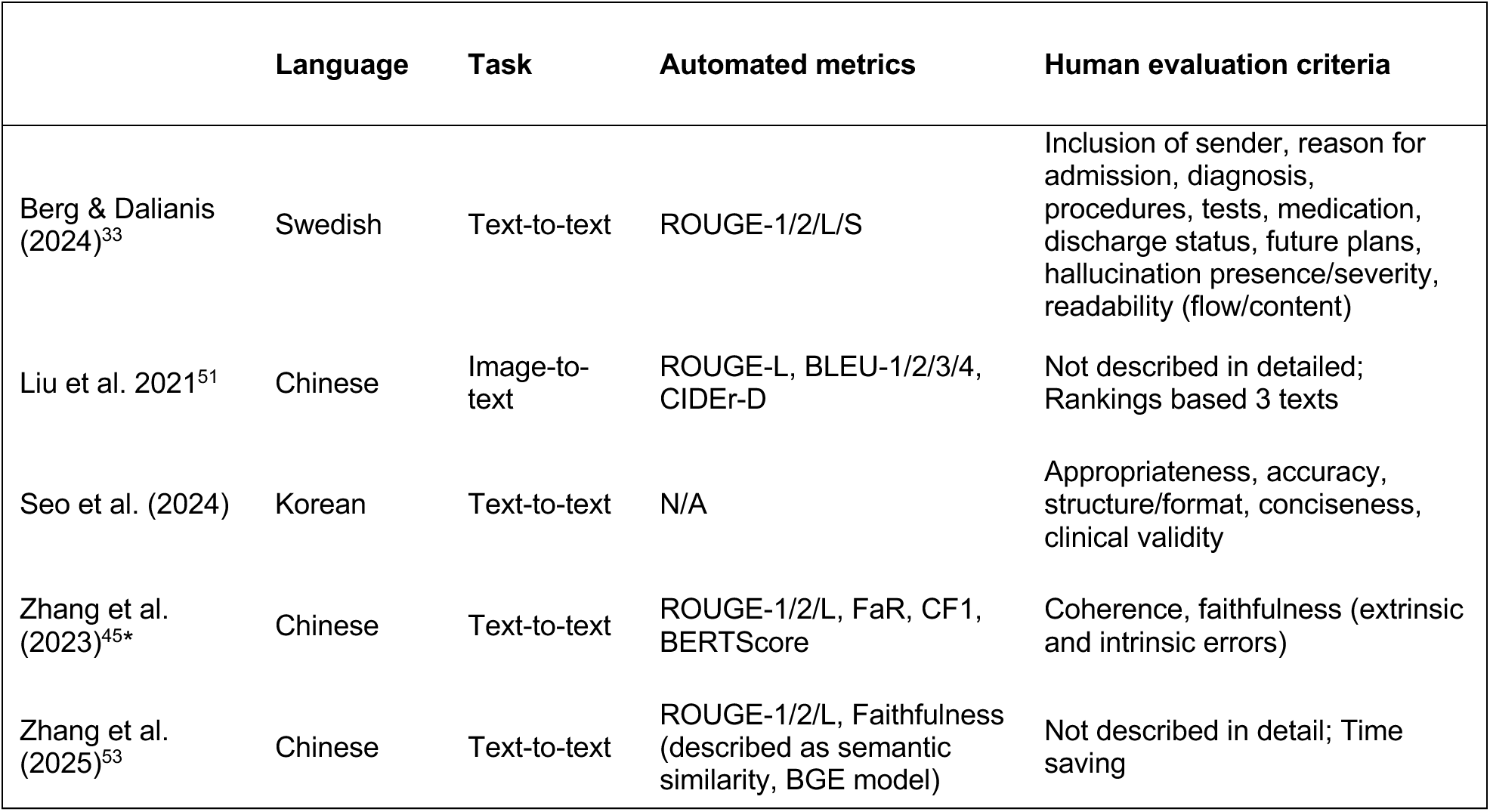
Non-English studies and reported evaluation approaches. The asterisk indicates that the non-English results were reported for only a dialogue subtask.

|  | Language | Task | Automated metrics | Human evaluation criteria |
| --- | --- | --- | --- | --- |
| Berg & Dalianis (2024)<sup>33</sup> | Swedish | Text-to-text | ROUGE-1/2/L/S | Inclusion of sender, reason for admission, diagnosis, procedures, tests, medication, discharge status, future plans, hallucination presence/severity, readability (flow/content) |
| Liu et al. 2021<sup>51</sup> | Chinese | Image-to-text | ROUGE-L, BLEU-1/2/3/4, CIDEr-D | Not described in detailed; Rankings based 3 texts |
| Seo et al. (2024) | Korean | Text-to-text | N/A | Appropriateness, accuracy, structure/format, conciseness, clinical validity |
| Zhang et al. (2023)<sup>45*</sup> | Chinese | Text-to-text | ROUGE-1/2/L, FaR, CF1, BERTScore | Coherence, faithfulness (extrinsic and intrinsic errors) |
| Zhang et al. (2025)<sup>53</sup> | Chinese | Text-to-text | ROUGE-1/2/L, Faithfulness (described as semantic similarity, BGE model) | Not described in detail; Time saving |

**Supplementary Table S9.** Fixed effect of perturbation severity from linear mixed models across metrics and models. Rows list metric *families* (Lexical overlap; Semantic/embedding; LLM evaluator), the specific *model*, *language*, and *edit type* (Delete, Modify, Paraphrase). The reported coefficient β_level (95% CI) is the fixed effect for a one-step increase in perturbation level (1→2→3) from models of the form *metric* ~ level + (1|case); p values are two-sided Wald tests. For similarity metrics (e.g., BLEU/ROUGE, BERTScore, BLEURT, cosine), more negative β indicates larger score decreases with more severe edits; for LLM change-score evaluators, more positive β indicates larger increases with severity. Cosine similarity uses OpenAI’s text-embedding-3-large embedding model. BH-adjusted *q* values and full outputs are provided in Supplementary data.

| Family | Model | Language | Edit type | $\beta_{\text{level}}$ (95% CI) | p |
| --- | --- | --- | --- | --- | --- |
| Lexical overlap | BLEU SCORE | English | Delete | -0.083 (-0.101 to -0.065) | <0.001 |
| Lexical overlap | BLEU SCORE | English | Modify | -0.114 (-0.158 to -0.070) | <0.001 |
| Lexical overlap | BLEU SCORE | English | Paraphrase | -0.048 (-0.054 to -0.043) | <0.001 |
| Lexical overlap | BLEU SCORE | Finnish | Delete | -0.092 (-0.107 to -0.077) | <0.001 |
| Lexical overlap | BLEU SCORE | Finnish | Modify | -0.113 (-0.155 to -0.071) | <0.001 |
| Lexical overlap | BLEU SCORE | Finnish | Paraphrase | -0.045 (-0.048 to -0.043) | <0.001 |
| Lexical overlap | BLEU SCORE | Swedish | Delete | -0.086 (-0.104 to -0.068) | <0.001 |
| Lexical overlap | BLEU SCORE | Swedish | Modify | -0.154 (-0.234 to -0.074) | <0.001 |
| Lexical overlap | BLEU SCORE | Swedish | Paraphrase | -0.042 (-0.048 to -0.036) | <0.001 |
| Lexical overlap | ROUGE 1 | English | Delete | -0.048 (-0.061 to -0.035) | <0.001 |
| Lexical overlap | ROUGE 1 | English | Modify | -0.079 (-0.117 to -0.042) | <0.001 |
| Lexical overlap | ROUGE 1 | English | Paraphrase | -0.020 (-0.022 to -0.018) | <0.001 |
| Lexical overlap | ROUGE 1 | Finnish | Delete | -0.057 (-0.069 to -0.044) | <0.001 |
| Lexical overlap | ROUGE 1 | Finnish | Modify | -0.087 (-0.127 to -0.047) | <0.001 |
| Lexical overlap | ROUGE 1 | Finnish | Paraphrase | -0.023 (-0.026 to -0.020) | <0.001 |
| Lexical overlap | ROUGE 1 | Swedish | Delete | -0.050 (-0.063 to -0.037) | <0.001 |
| Lexical overlap | ROUGE 1 | Swedish | Modify | -0.100 (-0.154 to -0.046) | <0.001 |
| Lexical overlap | ROUGE 1 | Swedish | Paraphrase | -0.022 (-0.026 to -0.017) | <0.001 |
| Lexical overlap | ROUGE 2 | English | Delete | -0.059 (-0.074 to -0.045) | <0.001 |
| Lexical overlap | ROUGE 2 | English | Modify | -0.112 (-0.160 to -0.065) | <0.001 |
| Lexical overlap | ROUGE 2 | English | Paraphrase | -0.046 (-0.050 to -0.043) | <0.001 |
| Lexical overlap | ROUGE 2 | Finnish | Delete | -0.072 (-0.085 to -0.059) | <0.001 |
| Lexical overlap | ROUGE 2 | Finnish | Modify | -0.118 (-0.170 to -0.066) | <0.001 |
| Lexical overlap | ROUGE 2 | Finnish | Paraphrase | -0.045 (-0.049 to -0.040) | <0.001 |
| Lexical overlap | ROUGE 2 | Swedish | Delete | -0.063 (-0.077 to -0.049) | <0.001 |
| Lexical overlap | ROUGE 2 | Swedish | Modify | -0.126 (-0.189 to -0.063) | <0.001 |
| Lexical overlap | ROUGE 2 | Swedish | Paraphrase | -0.040 (-0.047 to -0.033) | <0.001 |
| Lexical overlap | ROUGE 3 | English | Delete | -0.072 (-0.089 to -0.055) | <0.001 |
| Lexical overlap | ROUGE 3 | English | Modify | -0.127 (-0.177 to -0.077) | <0.001 |
| Lexical overlap | ROUGE 3 | English | Paraphrase | -0.065 (-0.072 to -0.058) | <0.001 |
| Lexical overlap | ROUGE 3 | Finnish | Delete | -0.087 (-0.101 to -0.072) | <0.001 |
| Lexical overlap | ROUGE 3 | Finnish | Modify | -0.133 (-0.189 to -0.077) | <0.001 |
| Lexical overlap | ROUGE 3 | Finnish | Paraphrase | -0.061 (-0.065 to -0.057) | <0.001 |
| Lexical overlap | ROUGE 3 | Swedish | Delete | -0.076 (-0.091 to -0.060) | <0.001 |
| Lexical overlap | ROUGE 3 | Swedish | Modify | -0.138 (-0.205 to -0.071) | <0.001 |
| Lexical overlap | ROUGE 3 | Swedish | Paraphrase | -0.055 (-0.064 to -0.045) | <0.001 |
| Lexical overlap | ROUGE 4 | English | Delete | -0.083 (-0.103 to -0.064) | <0.001 |
| Lexical overlap | ROUGE 4 | English | Modify | -0.139 (-0.192 to -0.086) | <0.001 |
| Lexical overlap | ROUGE 4 | English | Paraphrase | -0.081 (-0.090 to -0.072) | <0.001 |
| Lexical overlap | ROUGE 4 | Finnish | Delete | -0.099 (-0.115 to -0.084) | <0.001 |
| Lexical overlap | ROUGE 4 | Finnish | Modify | -0.144 (-0.203 to -0.085) | <0.001 |
| Lexical overlap | ROUGE 4 | Finnish | Paraphrase | -0.075 (-0.079 to -0.071) | <0.001 |
| Lexical overlap | ROUGE 4 | Swedish | Delete | -0.087 (-0.104 to -0.070) | <0.001 |
| Lexical overlap | ROUGE 4 | Swedish | Modify | -0.146 (-0.215 to -0.076) | <0.001 |
| Lexical overlap | ROUGE 4 | Swedish | Paraphrase | -0.066 (-0.079 to -0.054) | <0.001 |
| Semantic | bertscore f1 xlm-roberta-base | English | Delete | -0.015 (-0.018 to -0.011) | <0.001 |
| Semantic | bertscore f1 xlm-roberta-base | English | Modify | -0.019 (-0.029 to -0.009) | <0.001 |
| Semantic | bertscore f1 xlm-roberta-base | English | Paraphrase | -0.004 (-0.006 to -0.003) | <0.001 |
| Semantic | bertscore f1 xlm-roberta-base | Finnish | Delete | -0.019 (-0.024 to -0.014) | <0.001 |
| Semantic | bertscore f1 xlm-roberta-base | Finnish | Modify | -0.015 (-0.023 to -0.006) | <0.001 |
| Semantic | bertscore f1 xlm-roberta-base | Finnish | Paraphrase | -0.005 (-0.007 to -0.003) | <0.001 |
| Semantic | bertscore f1 xlm-roberta-base | Swedish | Delete | -0.015 (-0.019 to -0.011) | <0.001 |
| Semantic | bertscore f1 xlm-roberta-base | Swedish | Modify | -0.022 (-0.032 to -0.013) | <0.001 |
| Semantic | bertscore f1 xlm-roberta-base | Swedish | Paraphrase | -0.005 (-0.008 to -0.002) | 0.002 |
| Semantic | BLEURT SCORE | English | Delete | -0.087 (-0.152 to -0.021) | 0.009 |
| Semantic | BLEURT SCORE | English | Modify | -0.220 (-0.372 to -0.067) | 0.005 |
| Semantic | BLEURT SCORE | English | Paraphrase | -0.027 (-0.054 to -0.000) | 0.050 |
| Semantic | BLEURT SCORE | Finnish | Delete | 0.020 (-0.011 to 0.052) | 0.21 |
| Semantic | BLEURT SCORE | Finnish | Modify | 0.033 (-0.036 to 0.102) | 0.35 |
| Semantic | BLEURT SCORE | Finnish | Paraphrase | -0.000 (-0.000 to 0.000) | 1.00 |
| Semantic | BLEURT SCORE | Swedish | Delete | -0.100 (-0.183 to -0.018) | 0.017 |
| Semantic | BLEURT SCORE | Swedish | Modify | -0.050 (-0.121 to 0.021) | 0.17 |
| Semantic | BLEURT SCORE | Swedish | Paraphrase | -0.038 (-0.096 to 0.021) | 0.21 |
| Semantic | cosine similarity | English | Delete | -0.019 (-0.033 to -0.006) | 0.004 |
| Semantic | cosine similarity | English | Modify | -0.029 (-0.044 to -0.014) | <0.001 |
| Semantic | cosine similarity | English | Paraphrase | -0.002 (-0.003 to -0.001) | <0.001 |
| Semantic | cosine similarity | Finnish | Delete | -0.026 (-0.039 to -0.012) | <0.001 |
| Semantic | cosine similarity | Finnish | Modify | -0.037 (-0.057 to -0.016) | <0.001 |
| Semantic | cosine similarity | Finnish | Paraphrase | -0.003 (-0.005 to -0.002) | <0.001 |
| Semantic | cosine similarity | Swedish | Delete | -0.025 (-0.041 to -0.009) | 0.002 |
| Semantic | cosine similarity | Swedish | Modify | -0.032 (-0.053 to -0.012) | 0.002 |
| Semantic | cosine similarity | Swedish | Paraphrase | -0.003 (-0.005 to -0.001) | 0.008 |
| LLM evaluator | DeepSeek R1 | English | Delete | 2.500 (1.789 to 3.211) | <0.001 |
| LLM evaluator | DeepSeek R1 | English | Modify | 4.300 (3.265 to 5.335) | <0.001 |
| LLM evaluator | DeepSeek R1 | English | Paraphrase | 0.100 (-0.272 to 0.472) | 0.60 |
| LLM evaluator | DeepSeek R1 | Finnish | Delete | 2.400 (1.003 to 3.797) | <0.001 |
| LLM evaluator | DeepSeek R1 | Finnish | Modify | 1.600 (-0.220 to 3.420) | 0.085 |
| LLM evaluator | DeepSeek R1 | Finnish | Paraphrase | 0.800 (0.206 to 1.394) | 0.008 |
| LLM evaluator | DeepSeek R1 | Swedish | Delete | 2.900 (1.574 to 4.226) | <0.001 |
| LLM evaluator | DeepSeek R1 | Swedish | Modify | 1.600 (0.433 to 2.767) | 0.007 |
| LLM evaluator | DeepSeek R1 | Swedish | Paraphrase | 0.700 (0.054 to 1.346) | 0.034 |
| LLM evaluator | Gemma3n E2B | English | Delete | 0.700 (0.265 to 1.135) | 0.002 |
| LLM evaluator | Gemma3n E2B | English | Modify | 0.900 (0.572 to 1.228) | <0.001 |
| LLM evaluator | Gemma3n E2B | English | Paraphrase | -0.900 (-3.593 to 1.793) | 0.51 |
| LLM evaluator | Gemma3n E2B | Finnish | Delete | 0.600 (0.201 to 0.999) | 0.003 |
| LLM evaluator | Gemma3n E2B | Finnish | Modify | 1.100 (0.658 to 1.542) | <0.001 |
| LLM evaluator | Gemma3n E2B | Finnish | Paraphrase | -1.100 (-3.221 to 1.021) | 0.31 |
| LLM evaluator | Gemma3n E2B | Swedish | Delete | 0.400 (0.128 to 0.672) | 0.004 |
| LLM evaluator | Gemma3n E2B | Swedish | Modify | 1.100 (0.697 to 1.503) | <0.001 |
| LLM evaluator | Gemma3n E2B | Swedish | Paraphrase | 0.400 (-0.033 to 0.833) | 0.070 |
| LLM evaluator | Gemma3n E4B | English | Delete | 1.300 (0.936 to 1.664) | <0.001 |
| LLM evaluator | Gemma3n E4B | English | Modify | 3.300 (2.515 to 4.085) | <0.001 |
| LLM evaluator | Gemma3n E4B | English | Paraphrase | 2.300 (1.307 to 3.293) | <0.001 |
| LLM evaluator | Gemma3n E4B | Finnish | Delete | 1.600 (0.892 to 2.308) | <0.001 |
| LLM evaluator | Gemma3n E4B | Finnish | Modify | 3.800 (3.283 to 4.317) | <0.001 |
| LLM evaluator | Gemma3n E4B | Finnish | Paraphrase | 3.300 (2.251 to 4.349) | <0.001 |
| LLM evaluator | Gemma3n E4B | Swedish | Delete | 1.200 (0.538 to 1.862) | <0.001 |
| LLM evaluator | Gemma3n E4B | Swedish | Modify | 3.600 (2.698 to 4.502) | <0.001 |
| LLM evaluator | Gemma3n E4B | Swedish | Paraphrase | 2.200 (1.317 to 3.083) | <0.001 |
| LLM evaluator | gpt-oss-20b | English | Delete | 5.000 (2.737 to 7.263) | <0.001 |
| LLM evaluator | gpt-oss-20b | English | Modify | 4.500 (3.153 to 5.847) | <0.001 |
| LLM evaluator | gpt-oss-20b | Finnish | Modify | nan (nan to nan) | — |
| LLM evaluator | gpt-oss-20b | Finnish | Paraphrase | 0.125 (-0.154 to 0.404) | 0.38 |
| LLM evaluator | gpt-oss-20b | Swedish | Delete | 2.500 (1.934 to 3.066) | <0.001 |
| LLM evaluator | gpt-oss-20b | Swedish | Modify | 2.500 (-0.329 to 5.329) | 0.083 |
| LLM evaluator | gpt-oss-20b | Swedish | Paraphrase | 0.167 (-0.086 to 0.420) | 0.20 |
| LLM evaluator | gpt-oss-120b | English | Delete | 4.200 (3.211 to 5.189) | <0.001 |
| LLM evaluator | gpt-oss-120b | English | Modify | 8.500 (6.113 to 10.887) | <0.001 |
| LLM evaluator | gpt-oss-120b | Finnish | Delete | 4.100 (3.571 to 4.629) | <0.001 |
| LLM evaluator | gpt-oss-120b | Finnish | Modify | 7.400 (4.735 to 10.065) | <0.001 |
| LLM evaluator | gpt-oss-120b | Finnish | Paraphrase | 0.300 (0.083 to 0.517) | 0.007 |
| LLM evaluator | gpt-oss-120b | Swedish | Delete | 4.500 (3.417 to 5.583) | <0.001 |
| LLM evaluator | gpt-oss-120b | Swedish | Modify | 5.400 (2.745 to 8.055) | <0.001 |
| LLM evaluator | gpt-oss-120b | Swedish | Paraphrase | 0.100 (-0.056 to 0.256) | 0.21 |
| LLM evaluator | gpt-3.5 | English | Delete | 3.500 (2.117 to 4.883) | <0.001 |
| LLM evaluator | gpt-3.5 | English | Modify | 2.300 (0.857 to 3.743) | 0.002 |
| LLM evaluator | gpt-3.5 | English | Paraphrase | 0.200 (-0.725 to 1.125) | 0.67 |
| LLM evaluator | gpt-3.5 | Finnish | Delete | 1.500 (0.443 to 2.557) | 0.005 |
| LLM evaluator | gpt-3.5 | Finnish | Modify | 2.400 (0.634 to 4.166) | 0.008 |
| LLM evaluator | gpt-3.5 | Finnish | Paraphrase | 2.300 (0.986 to 3.614) | <0.001 |
| LLM evaluator | gpt-3.5 | Swedish | Delete | 1.900 (-0.544 to 4.344) | 0.13 |
| LLM evaluator | gpt-3.5 | Swedish | Modify | 3.600 (1.717 to 5.483) | <0.001 |
| LLM evaluator | gpt-3.5 | Swedish | Paraphrase | 0.700 (-0.242 to 1.642) | 0.15 |
| LLM evaluator | gpt-4 | English | Delete | 3.500 (2.292 to 4.708) | <0.001 |
| LLM evaluator | gpt-4 | English | Modify | 5.400 (3.908 to 6.892) | <0.001 |
| LLM evaluator | gpt-4 | Finnish | Delete | 2.900 (1.268 to 4.532) | <0.001 |
| LLM evaluator | gpt-4 | Finnish | Modify | 4.600 (3.746 to 5.454) | <0.001 |
| LLM evaluator | gpt-4 | Swedish | Delete | 3.900 (2.324 to 5.476) | <0.001 |
| LLM evaluator | gpt-4 | Swedish | Modify | 5.400 (3.777 to 7.023) | <0.001 |
| LLM evaluator | gpt-4.1 | English | Delete | 2.500 (1.820 to 3.180) | <0.001 |
| LLM evaluator | gpt-4.1 | English | Modify | 3.100 (2.381 to 3.819) | <0.001 |
| LLM evaluator | gpt-4.1 | English | Paraphrase | 0.100 (-0.256 to 0.456) | 0.58 |
| LLM evaluator | gpt-4.1 | Finnish | Delete | 2.100 (1.519 to 2.681) | <0.001 |
| LLM evaluator | gpt-4.1 | Finnish | Modify | 3.400 (2.378 to 4.422) | <0.001 |
| LLM evaluator | gpt-4.1 | Finnish | Paraphrase | 0.200 (-0.370 to 0.770) | 0.49 |
| LLM evaluator | gpt-4.1 | Swedish | Delete | 3.200 (2.371 to 4.029) | <0.001 |
| LLM evaluator | gpt-4.1 | Swedish | Modify | 2.400 (1.212 to 3.588) | <0.001 |
| LLM evaluator | gpt-4.1 | Swedish | Paraphrase | 0.300 (0.083 to 0.517) | 0.007 |
| LLM evaluator | gpt-4o | English | Delete | 3.400 (1.837 to 4.963) | <0.001 |
| LLM evaluator | gpt-4o | English | Modify | 3.900 (3.042 to 4.758) | <0.001 |
| LLM evaluator | gpt-4o | English | Paraphrase | 0.300 (-0.167 to 0.767) | 0.21 |
| LLM evaluator | gpt-4o | Finnish | Delete | 4.300 (1.805 to 6.795) | <0.001 |
| LLM evaluator | gpt-4o | Finnish | Modify | 3.800 (2.933 to 4.667) | <0.001 |
| LLM evaluator | gpt-4o | Finnish | Paraphrase | 0.400 (-0.175 to 0.975) | 0.17 |
| LLM evaluator | gpt-4o | Swedish | Delete | 3.700 (1.955 to 5.445) | <0.001 |
| LLM evaluator | gpt-4o | Swedish | Modify | 4.300 (1.758 to 6.842) | <0.001 |
| LLM evaluator | gpt-5 mini | English | Delete | 4.800 (4.076 to 5.524) | <0.001 |
| LLM evaluator | gpt-5 mini | English | Modify | 5.400 (4.312 to 6.488) | <0.001 |
| LLM evaluator | gpt-5 mini | English | Paraphrase | 0.100 (-0.056 to 0.256) | 0.21 |
| LLM evaluator | gpt-5 mini | Finnish | Delete | 4.500 (3.447 to 5.553) | <0.001 |
| LLM evaluator | gpt-5 mini | Finnish | Modify | 5.400 (3.808 to 6.992) | <0.001 |
| LLM evaluator | gpt-5 mini | Finnish | Paraphrase | 0.100 (-0.124 to 0.324) | 0.38 |
| LLM evaluator | gpt-5 mini | Swedish | Delete | 4.900 (4.228 to 5.572) | <0.001 |
| LLM evaluator | gpt-5 mini | Swedish | Modify | 5.200 (3.586 to 6.814) | <0.001 |
| LLM evaluator | gpt-5 mini | Swedish | Paraphrase | 0.100 (-0.056 to 0.256) | 0.21 |
| LLM evaluator | gpt-5 nano | English | Delete | 3.300 (2.173 to 4.427) | <0.001 |
| LLM evaluator | gpt-5 nano | English | Modify | 5.600 (4.004 to 7.196) | <0.001 |
| LLM evaluator | gpt-5 nano | English | Paraphrase | 0.100 (-0.272 to 0.472) | 0.60 |
| LLM evaluator | gpt-5 nano | Finnish | Delete | 3.400 (1.690 to 5.110) | <0.001 |
| LLM evaluator | gpt-5 nano | Finnish | Modify | 5.100 (3.976 to 6.224) | <0.001 |
| LLM evaluator | gpt-5 nano | Finnish | Paraphrase | 0.100 (-0.129 to 0.329) | 0.39 |
| LLM evaluator | gpt-5 nano | Swedish | Delete | 4.600 (3.698 to 5.502) | <0.001 |
| LLM evaluator | gpt-5 nano | Swedish | Modify | 5.000 (4.347 to 5.653) | <0.001 |
| LLM evaluator | gpt-5 nano | Swedish | Paraphrase | -0.100 (-0.431 to 0.231) | 0.55 |

